# Resistive Load During CPAP and Automatic Tube Compensation (ATC): A Bench Comparison of ICU Ventilators

**DOI:** 10.64898/2026.07.08.26357537

**Authors:** Ben Fabry, Christian Kuster, Roland C. E. Francis

**Affiliations:** Department of Physics, Friedrich-Alexander Universität Erlangen-Nürnberg, Erlangen, Germany; Department of Anesthesiology, Universitätsklinikum Erlangen, Friedrich-Alexander Universität Erlangen-Nürnberg, Erlangen, Germany

**Author notes:** Corresponding author:* Ben Fabry Department of Physics Friedrich-Alexander Universität Erlangen-Nürnberg Henkestr. 91 91052 Erlangen Germany.

**Keywords:** patient-ventilator asynchrony, imposed work of breathing, closed-loop pressure control, demand-flow control

## Abstract

**Background:** Automatic tube compensation (ATC) aims to compensate for the additional resistive load imposed by the endotracheal tube during spontaneous breathing. In ATC mode, the ventilator adds the flow-dependent pressure drop across the tube during inspiration and subtracts it during expiration so that tracheal pressure remains near PEEP. Clinical studies with a prototype ventilator demonstrated clear physiological benefits of ATC, but subsequent studies with commercial systems have not confirmed these findings. A 2003 bench study found that commercial ventilators provided, at best, only partial tube compensation that is unlikely to result in meaningful clinical benefits. We therefore tested the tube compensation performance in contemporary ICU ventilators.

**Methods:** We performed a bench comparison of six commercial ICU ventilators and an ATC prototype ventilator. An active lung simulator generated spontaneous breathing patterns with weak, moderate, and strong inspiratory efforts at different PEEP levels. We tested endotracheal tubes with inner diameters of 7 and 8 mm, and measured airway pressure, tracheal pressure, and flow during CPAP with and without ATC. Breathing through the tube against open atmosphere served as a zero-PEEP/T-piece reference.

**Results:** In CPAP mode, commercial ventilators showed substantial flow-dependent airway-pressure deviations, amounting to an added resistance of 0.7 - 6.5 mbar·s/L, whereas the resistance imposed by the ATC prototype never exceeded 0.6 mbar·s/L. Averaged across all conditions with PEEP > 0 mbar, commercial ventilators in ATC mode reduced the resistive load relative to T-piece breathing by no more than 27%, while some models even imposed an additional load. By contrast, the ATC prototype maintained tracheal pressure close to PEEP throughout the breathing cycle and eliminated 79% of the load on average.

**Conclusions:** The commercial ventilators tested in this study failed to provide effective tube compensation despite a nominal 100% ATC setting. More advanced approaches, such as those implemented in the ATC prototype, may be required for stable and physiologically meaningful tracheal-pressure control.

## Introduction

The endotracheal tube imposes a major additional, nonlinear, flow-dependent resistive load in intubated patients. At a flow of 1 L/s, the pressure drop across a clean 8-mm-ID tube is approximately 6.5 mbar (∼ 6.6 cm H_2O_), and 10.5 mbar (∼ 10.7 cmH_2O_) across a 7-mm-ID tube [1, 2]. During controlled mechanical ventilation, this resistance impedes airflow and can cause dynamic hyperinflation and intrinsic PEEP [3, 4]. During ventilator-assisted spontaneous breathing, the endotracheal tube can dramatically increase the work of breathing [5] and promote patient-ventilator asynchrony [6]. During strong inspiratory efforts, it can also cause large negative swings in tracheal and alveolar pressure [5] that have been implicated in patient self-inflicted lung injury [7]. The imposed tube resistive load can mimic weaning intolerance and delay extubation [8].

Automatic tube compensation (ATC) was developed to remove the artificial resistance of the endotracheal tube and prevent tube-related complications during mechanical ventilation [9]. The principle of ATC is straightforward: the ventilator continuously estimates the pressure drop across the endotracheal tube from the measured flow and the known pressure-flow characteristics of the tube. It then dynamically adds this pressure during inspiration and subtracts it during expiration, so that the tracheal pressure remains close to the set PEEP level at all times, regardless of the magnitude of inspiratory effort and flow [9]. Thus, ATC applies a variable, continuously flow-adapted positive airway pressure during inspiration and a variable sub-PEEP pressure during expiration. The physiological goal is to make the endotracheal tube mechanically “invisible” to the patient.

Early clinical studies with an ATC prototype ventilator established that the concept was physiologically sound and clinically beneficial. ATC compensated tube-related work of breathing in critically ill patients with high ventilatory demand, where conventional pressure support of 5, 10, or 15 mbar failed to do so [5]. Under ATC, the tracheal pressure remained close to PEEP even during strong respiratory efforts when the pressure loss across the tube exceeded 25 mbar [5]. In a patient with severe patient-ventilator desynchronization under conventional, triggered inspiratory pressure support ventilation, the combination of ATC and proportional assist ventilation restored breath-by-breath support even at respiratory rates of more than 50 breaths per minute [10].

In 2003, a bench study compared the original ATC prototype with commercial ATC implementations [11], and found that commercial systems reduced the tube-related work of breathing by only 50-68% during inspiration, whereas the original ATC prototype achieved a reduction by 97%. During expiration, commercial systems achieved reductions of tube-related work by only 1-26%, whereas the original ATC prototype reduced it by 70%. The authors concluded that, unless these systems were improved, the advantages of commercial ATC over conventional patient-triggered pressure support would probably be negligible.

Here, we tested whether contemporary commercial ICU ventilators in ATC mode achieve the physiological goal of maintaining near-constant tracheal pressure during spontaneous breathing at different levels of respiratory effort. We compared four contemporary commercial ventilators with two previous-generation commercial ventilators and with an ATC prototype during simulated spontaneous breathing through endotracheal tubes of different sizes.

## Methods

ATC can be provided either as a stand-alone mode or as an add-on feature to other modes such as patient-triggered pressure support or proportional assist ventilation. In the present study, we only consider ATC as a stand-alone mode, not its combination with other modes.

We tested the following ventilators, all of which offer Automatic Tube Compensation (ATC), also called Tube Resistance Compensation (TRC): Dräger Evita 4, Dräger Evita XL, Dräger Evita V600 (Drägerwerk AG, Lübeck, Germany), Löwenstein Elisa 800 (Löwenstein Medical, Bad Ems, Germany), Hamilton C2, Hamilton C6 (Hamilton Medical AG, Bonaduz, Switzerland), and an ATC prototype ventilator. The original ATC prototype used in the clinical studies of the 1990s [5, 6, 9, 10] and in the 2003 technical comparison study [11] was no longer available. In this study, we used an ATC prototype based on the system described previously [9], with the following modifications (see Fig. S4). Positive and negative pressure were generated by two radial blowers (U65HN-024KS-6, Micronel, Switzerland), which were connected to the inspiratory and expiratory limbs of a Y-piece. Pressure was controlled by a three-way valve integrated directly into the Y-piece, as described in [12]. The prototype ventilator was not certified for patient-use. We did not apply any gas blending, only room air was used. No humidification or heating was applied, and no bacterial or HME filters were added.

We tested tube-resistance compensation with original-length endotracheal tubes (Rüsch Super Safety Clear, Teleflex, Ireland) with inner diameters of 7 and 8 mm. Each tube was inserted into an artificial trachea made of polymethyl methacrylate (PMMA) and sealed airtight with the cuff. The artificial trachea had an inner diameter of 2.1 cm. Tracheal pressure was measured through 12 small radial holes equally spaced around the circumference of the artificial trachea and connected to a piezoresistive differential pressure sensor (Honeywell HCS series, USA; range ±80 mbar). The tracheal pressure measurement site was located 60 mm distal to the tube tip and 50 mm proximal to the distal end of the artificial trachea, as described previously [1, 4].

On the proximal side, the endotracheal tube was connected to a swivel connector, a flexible tube, a thermal mass flow sensor (SFM3300, Sensirion, Switzerland), a short tube with a ring channel with eight small radial holes for airway pressure measurement, and the Y-piece of the ventilator under test. Airway pressure was measured with a piezoresistive differential pressure sensor (Honeywell HCS series, USA; range ±80 mbar). Pressure and flow signals were sampled at 250 Hz and low-pass filtered using a zero-phase 4th-order Bessel filter with a cutoff frequency of 25 Hz.

On the distal side, the artificial trachea was connected to a custom-built active lung simulator, as described in detail previously [4]. The simulator consisted of a 1.8 L piston pump connected to a 20 L glass container loosely filled with steel wool. A stepper-motor drive controlled the piston motion so that the simulator reproduced a prescribed airway resistance, respiratory-system elastance, and time-varying muscle pressure. The maximum response frequency of the active lung simulator was 25 Hz. The 20 L glass container acted as a compliance reservoir and prevented pressure and flow components above this frequency from encountering an effectively stiff simulator. This resulted in a minimum compliance of 20 ml/mbar, independent of the response bandwidth of the lung simulator.

We tested three different breathing patterns, corresponding to weak, moderate, and strong inspiratory efforts and increasing respiratory demand. For weak and moderate effort, airway resistance was set to 2 mbar·s/L. Inspiratory time t_in_ was set to 1 s, and expiratory time to 2 s. The minimum inspiratory muscle pressure, P_mus,min_, was set to −5 mbar for weak effort and −10 mbar for moderate effort. During inspiration, P_mus_ decreased monotonically from zero to P_mus,min_ according to a quarter sine wave, P_mus_(t) = P_mus,min_ sin[π t / (2 t_in_)], where t denotes the time after the onset of inspiration. During expiration, P_mus_ returned to zero within 0.5 s according to a half sine wave, P_mus_(t) = 0.5 P_mus,min_ [1 + cos(π (t - t_in_) / 0.5 s)]. Total respiratory compliance was set to 100 ml/mbar for weak effort, and 50 ml/mbar for moderate effort. This lower compliance during moderate effort reflects the reduced chest-wall compliance that occurs during respiratory muscle activation. For simplicity, compliance was kept constant throughout the breathing cycle and was not varied with instantaneous muscle activity.

For strong inspiratory effort, the piston pump generated a sinusoidal volume pattern with a peak flow of 1 L/s, an inspiratory time of 1 s, and an expiratory time of 1 s, independent of the resulting pressures and flows at the airway. Thus, no muscle pressure or airway resistance was prescribed for this condition. The effective compliance was determined by the 20 L glass container and was approximately 20 ml/mbar, which corresponds to a relatively stiff respiratory system during strong inspiratory effort. The purpose of prescribing piston-movement rather than P_mus_ was to generate comparable flow conditions that are largely independent of the added load by the tube and the ventilator circuit.

We tested all ventilators both in CPAP and ATC modes. In CPAP mode, airway pressure should remain close to the set PEEP level throughout the breathing cycle, independent of respiratory effort and flow. In ATC mode, tracheal pressure should remain close to the set PEEP level. In the ventilator setup menu, we selected a tube diameter of 7 mm or 8 mm, depending on the inner diameter of the endotracheal tube used in each experiment, with 100% compensation in inspiration and - if possible - in expiration. The compensation algorithms and resistance parameters used by the tested ventilators were not consistently disclosed by the manufacturers. For the Dräger ventilators, the manual specifies a purely quadratic tube model, Δp_ETT_ = *k*_2_ |V̇|V̇, where V̇ is the flow through the tube. For full-length 8 mm and 7 mm tubes, respectively, the tabulated k_2_ values in the Evita 4 and Evita XL are 6.57 and 10.56 mbar/(L/s)^2^. The corresponding values for the Evita V600 are 4.54 and 8.08 mbar/(L/s)^2^. For the other commercial ventilators tested here, the exact equations and tube-resistance parameters were, to our knowledge, not publicly disclosed. The equation for controlling ATC in the prototype ventilator was 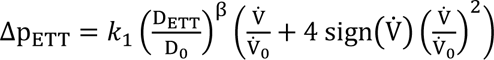, with k_1_ = 0.72 mbar, D_0_ = 10 mm, V̇_0_= 1 L/s, and exponent β = −3.6 [4]. D_ETT_ is the inner diameter of the endotracheal tube.

To quantify the pressure-control performance of the tested ventilators in CPAP mode, we computed the standard deviation of the airway pressure signal over the breathing cycle and normalized it by the standard deviation of flow, R_eff,aw_ = σ(P_aw_)/ σ(V̇). This ratio has units of resistance and in CPAP mode quantifies the effective resistance R_eff,aw_ proximal to the airway pressure measuring site, hence the resistance added by the ventilator circuit, demand-flow valve, and expiratory valve. Thus, R_eff,aw_ is a pragmatic measure of pressure-control performance under dynamic conditions; it does not distinguish true resistive pressure losses from inertive, compliant, or control-transient contributions. Because it is based on standard deviations across the breathing cycle, it is insensitive to the absolute PEEP level, to the sign of pressure deviations from PEEP, and to the timing of these deviations within the breathing cycle.

Analogous to R_eff,aw_, we also calculated the effective resistance at the tracheal pressure measurement site, R_eff,trach_, as the ratio of the standard deviation of tracheal pressure to the standard deviation of flow. In CPAP and ATC modes, R_eff,trach_ reflects the net effective resistive load of the combined endotracheal tube-ventilator system, which includes contributions from the swivel connector, flexible tubing, flow sensor, ventilator circuit, demand-flow valve, and expiratory valve.

We also calculated an inspiration-only effective tracheal resistance, R_eff,trach,in_, over the simulator-defined inspiratory-effort interval as the mean tracheal pressure deficit below intrinsic PEEP (iPEEP) divided by the mean positive inspiratory flow over the same interval. Throughout this interval, only pressure below iPEEP contributed to the mean pressure numerator, and only positive flow contributed to the inspired-volume denominator. iPEEP was approximated by the tracheal pressure at the simulator-defined end of expiration. This approximation was exact in the strong-effort condition because the airway resistance of the lung-simulator was set to zero. In the low- and moderate-effort conditions, it was a close approximation because end-expiratory flow, and hence the resistive pressure loss, was nearly zero. R_eff,trach,in_ is expressed in mbar·s/L and is algebraically and numerically identical to the volume-normalized inspiratory tracheal pressure-time product (PTP).

In ATC mode, we also report the additional work of breathing (WOB_add_), calculated over the same simulator-defined inspiratory-effort interval by integrating the tracheal pressure deficit below iPEEP with respect to positive inspired volume and normalizing it to the total positive inspired volume over that interval [5]. Pressure above iPEEP and during expiratory flow contributed zero. WOB_add_ is expressed in J/L.

We also measured the inspiratory and expiratory flow delays, defined as the intervals from the simulator-defined onset of inspiration and expiration, respectively, to the corresponding measured-flow transition. The reported mean flow-transition delay was the arithmetic mean of these two delays for each breath.

For all ventilators, breathing patterns, and tube diameters, we measured R_eff,trach_, PTP and WOB_add_ at PEEP levels of 0, 5, and 10 mbar. Moderate and strong effort were also tested at 15 mbar PEEP. The weak effort situation with a compliance of 100 ml/mbar could not be measured at 15 mbar PEEP as the resulting lung volume would have exceeded the 1.8 L capacity of the lung simulator. For each breathing pattern and tube diameter, we also measured R_eff,trach_, PTP and WOB_add_ during breathing against open atmosphere, with the ventilator disconnected. This condition corresponds to a zero-PEEP T-piece trial. In this case, R_eff,trach_ reflects the effective resistance of the endotracheal tube, swivel connector, flexible tubing, and sensors, without the added resistance of the ventilator valves and breathing circuit, and without resistance reduction by tube compensation.

In preliminary measurements, we observed that the pressure-control performance of the Evita V600 improved after approximately 1 min of simulated breathing, possibly reflecting adaptation of the ventilator’s control algorithm to the respiratory mechanics of the lung simulator. We therefore allowed all ventilators to operate for at least 1 min and began recording only once the pressure waveforms had stabilized. We analyzed 10 ± 3 breaths per condition. Results for each condition are reported as mean ± SEM across all analyzed breaths (Table S1). We also quantified the efficiency of tube compensation as the percentage reduction in R_eff,trach_, PTP, or WOB_add_ under ATC relative to its corresponding reference value measured either during the T-piece trial (Table S2) or during CPAP with the same ventilator (Table S3): compensation (%) = 100 × [1 - (ATC value/reference value)].

## Results

### Airway pressure control performance in CPAP mode

We first tested airway-pressure control during CPAP. In this mode, airway pressure should remain close to the set PEEP throughout the breathing cycle. However, all commercial ventilators showed substantial flow-dependent deviations of airway pressure from PEEP (Fig. 1). These deviations increased with inspiratory effort and occurred during both inspiration and expiration. In the Hamilton ventilators, peak deviations exceeded 3 mbar under strong effort. Thus, even in CPAP mode, before tube compensation is considered, commercial ventilators behave as non-ideal pressure sources because of limitations in closed-loop pressure control and valve dynamics. By contrast, the ATC prototype ventilator controlled airway pressure close to the target and exhibited noticeable deviations only during transitions between inspiration and expiration.

**Fig. 1:**
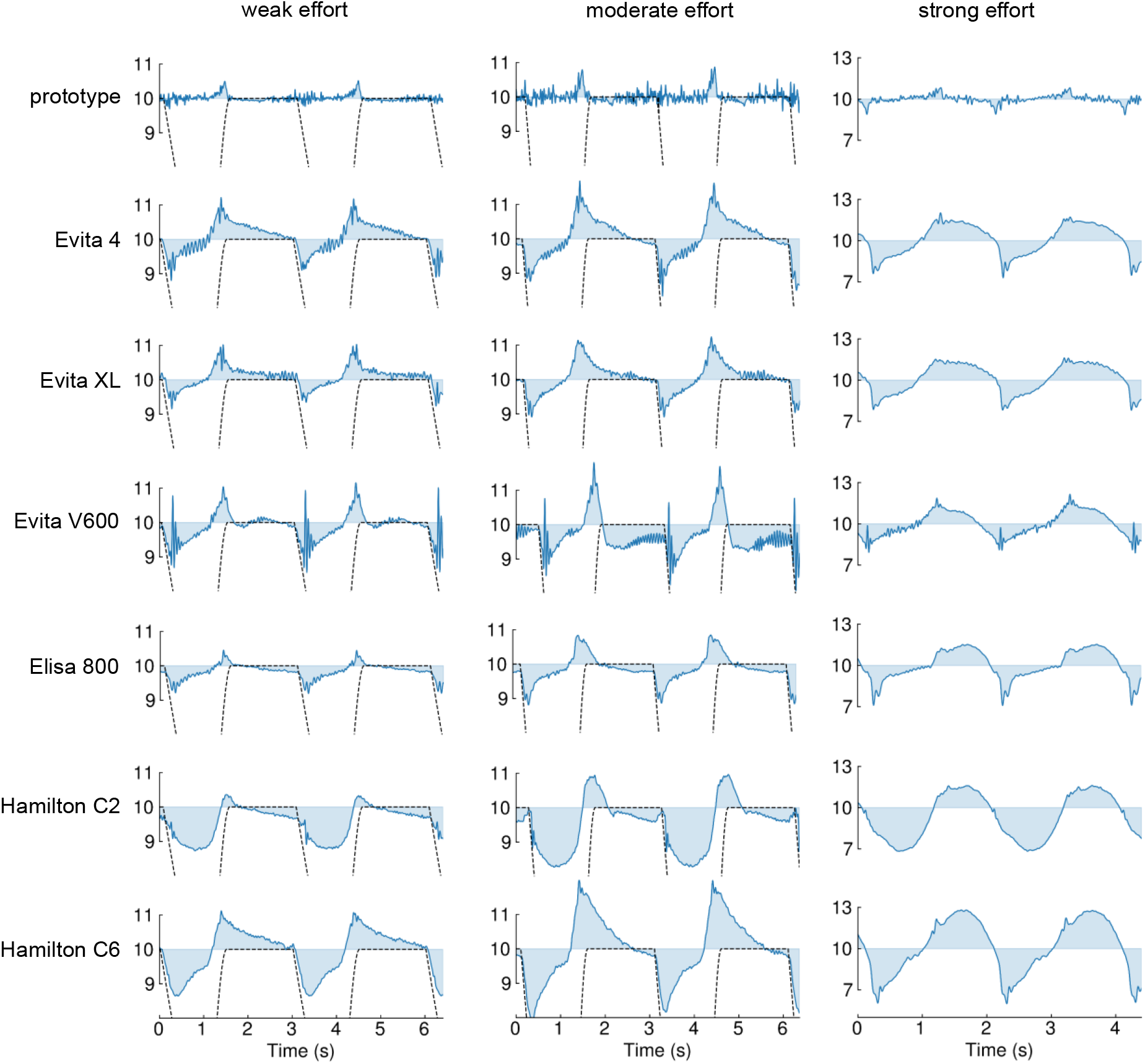
Airway pressure control during CPAP. Representative airway-pressure recordings (in mbar) during simulated spontaneous breathing through a 7 mm endotracheal tube at PEEP = 10 mbar. Columns show weak, moderate, and strong inspiratory effort. Rows show the tested ventilators. Airway pressure (blue) is plotted during two consecutive breathing cycles; the horizontal line indicates the target pressure. In an ideal CPAP system, airway pressure would remain on this line throughout the breathing cycle. The dotted line shows P_mus_ + PEEP and marks the timing of inspiration. The P_mus_ trace is truncated to keep the pressure deviations visible; the full, untruncated P_mus_ trace is shown in Fig. 3. Commercial ventilators showed substantial flow-dependent pressure deviations that increased with respiratory effort.

### Effective resistance in CPAP mode

We quantified the effective resistance R_eff,aw_ imposed by the ventilator during CPAP, and found values between approximately 0.7 and 6.5 mbar·s/L for the commercial ventilators (Fig. 2). Because R_eff,aw_ normalizes pressure deviations by flow, the larger pressure deviations at stronger respiratory effort did not translate into a higher effective resistance. Across most commercial ventilators, R_eff,aw_ was larger at zero PEEP than at higher PEEP levels. This PEEP dependence was particularly pronounced in the Hamilton C6. By contrast, the effective resistance imposed by the ATC prototype ventilator remained at 0.5 mbar·s/L on average and showed no systematic PEEP dependence.

**Fig. 2:**
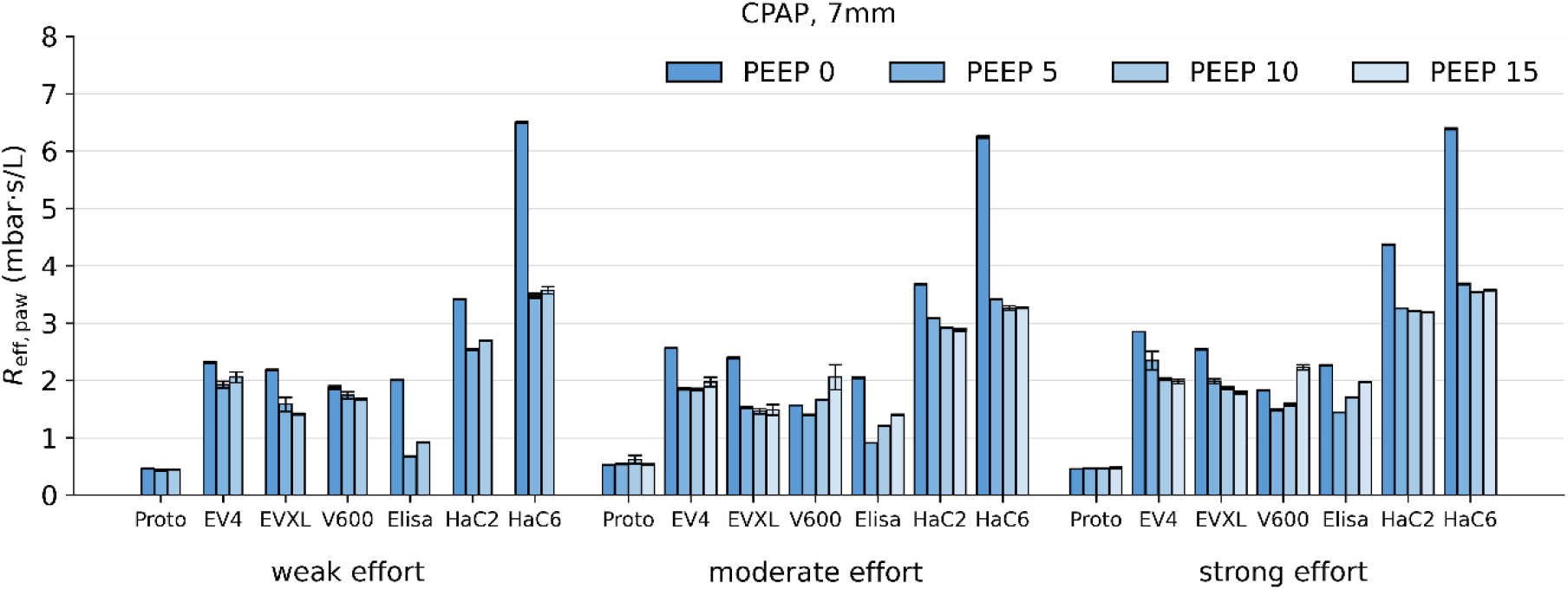
Effective airway resistance during CPAP. Effective airway resistance, R_eff,aw_, during CPAP with a 7 mm endotracheal tube. Values are shown for weak, moderate, and strong inspiratory effort and for different PEEP levels and ventilators. R_eff,aw_, during breathing in the “open” configuration, i.e. with the Y-piece removed and open to atmosphere, is zero by definition and not shown here. Values are mean ± SEM from 10 ± 3 breaths per condition. Corresponding effective airway resistance values during CPAP with an 8-mm tube were similar (see Table S1).

### Tracheal pressure control performance in ATC mode

Next, we tested tracheal-pressure control during Automatic Tube Compensation. In this mode, tracheal pressure should remain close to the set PEEP throughout the breathing cycle. Accordingly, airway pressure should increase above PEEP during inspiration and decrease below PEEP during expiration. This expected behavior was observed with the ATC prototype ventilator (Fig. 3). By contrast, all commercial ventilators showed substantial flow-dependent deviations of tracheal pressure from PEEP. These deviations increased with inspiratory effort and occurred during both inspiration and expiration. The two older Evita 4 and Evita XL ventilators did not offer expiratory tube compensation. Accordingly, airway pressure did not decrease below PEEP during expiration. However, both ventilators maintained nearly constant tracheal pressure during inspiration at weak and moderate effort. The Hamilton C6 also did not lower airway pressure below PEEP during expiration, even though expiratory tube compensation was activated. Taken together, the commercial ventilators did not maintain constant tracheal pressure throughout the breathing cycle as intended in ATC mode, either by design, as in the older Evita 4 and Evita XL models, or because tube compensation was insufficient.

**Fig. 3:**
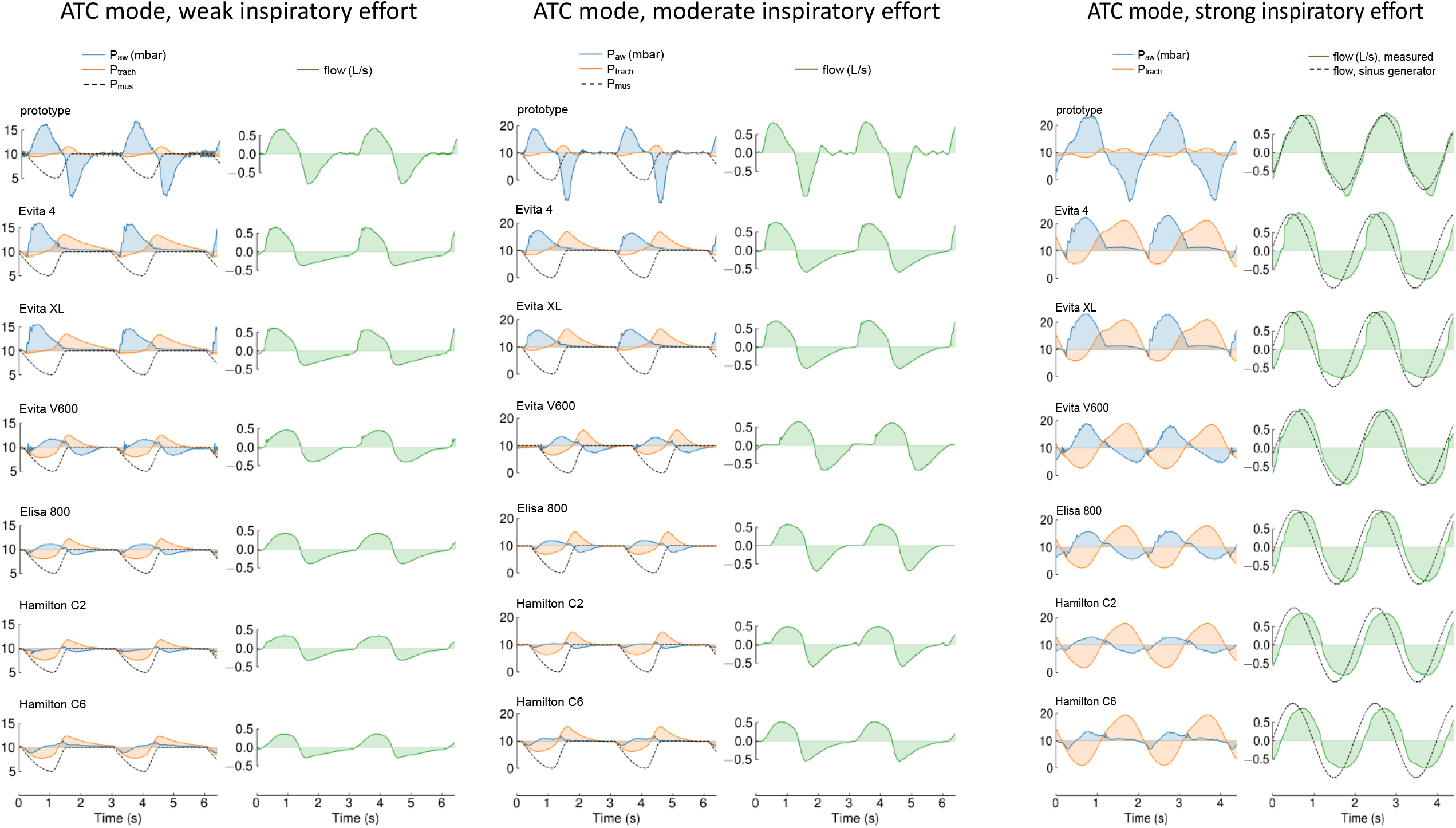
Tracheal-pressure control during ATC. Representative airway-pressure, tracheal-pressure, and muscle-pressure recordings during simulated spontaneous breathing through a 7 mm endotracheal tube at PEEP = 10 mbar. Separate panels show weak, moderate, and strong inspiratory effort. Airway pressure is shown in blue, tracheal pressure in orange, and muscle pressure plus PEEP as a dashed black line. For strong effort, the dashed line indicates the flow generated by the piston pump of the lung simulator. Pressures are plotted during two consecutive breathing cycles. The horizontal line at 10 mbar indicates the target pressure. In an ideal ATC system, tracheal pressure would remain on this line throughout the breathing cycle. The ATC prototype ventilator maintained tracheal pressure close to PEEP, whereas commercial ventilators showed substantial flow-dependent tracheal-pressure deviations that increased with inspiratory effort.

### Effective resistance in ATC mode

Next, we quantified the residual effective resistance R_eff,trach_ during ATC. The ATC prototype ventilator showed values below 2 mbar·s/L, without a pronounced dependence on inspiratory effort or PEEP (Fig. 4). By contrast, the commercial ventilators showed R_eff,trach_ values between approximately 4 and 13 mbar·s/L for a 7 mm tube, and between 3.3 and 11.5 mbar·s/L for an 8 mm tube. R_eff,trach_ increased markedly with stronger inspiratory effort. In the Evita V600, Elisa 800 and the two Hamilton ventilators, R_eff,trach_ was lower at higher PEEP levels than at zero PEEP, consistent with the ability of these ventilators to lower the airway pressure below PEEP during expiration and therefore to compensate expiratory tube resistance. The effective resistance during breathing against open atmosphere was not substantially higher than during ATC with any of the commercial ventilators. In some cases, commercial ATC imposed an even larger effective resistance than unsupported breathing through the tube against atmosphere.

**Fig. 4:**
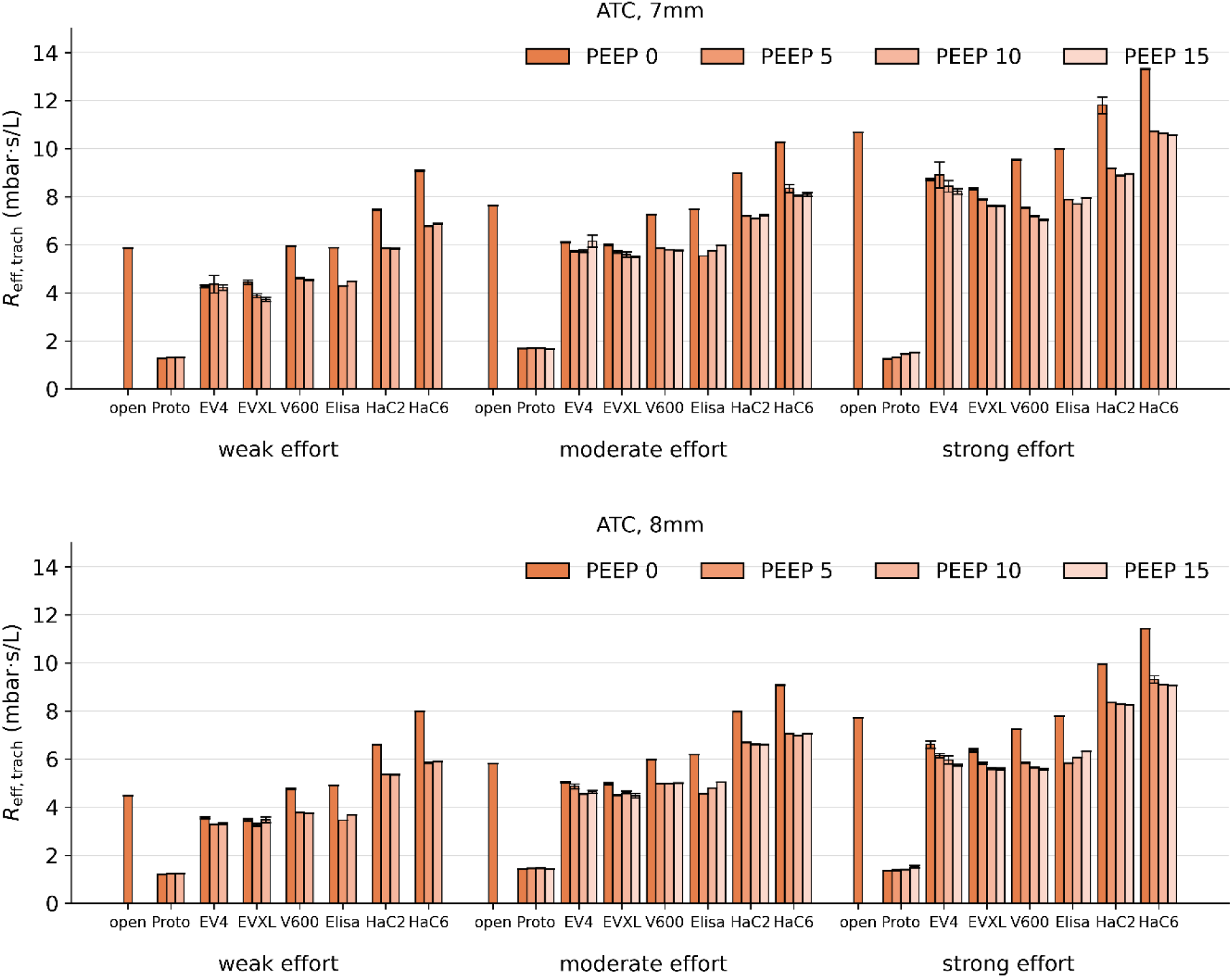
Effective tracheal resistance during ATC. Effective tracheal resistance, R_eff,trach_, during ATC with a 7 mm (top) or 8 mm (bottom) endotracheal tube, shown for weak, moderate, and strong inspiratory effort, different PEEP levels, and all tested ventilators. The “open” condition shows the effective resistance during breathing against open atmosphere with the ventilator disconnected, equivalent to a zero-PEEP T-piece trial. In an ideal ATC system, R_eff,trach_ should approach zero. Values are mean ± SEM from 10 ± 3 breaths per condition.

Averaged for all conditions, PEEP levels, and tube diameters, the Evita 4 in ATC mode compensated 22% of the effective tube resistance measured during unsupported breathing, the Evita XL compensated 25%, the Evita V600 compensated 17%, and the Elisa 800 compensated 16%. With the Hamilton C2 and C6, the effective resistance even increased by 9% and 24% relative to the effective tube resistance measured during unsupported breathing. Excluding PEEP 0, as it is rarely used in clinical practice, gave average compensation values of 23%, 27%, 23% and 23% for the Evita 4, XL, V600, and the Elisa 800, respectively. The Hamilton C2 and C6 still increased the load by 2% and 15%, respectively. Taken together, the commercial ventilators did not provide an effective tube-resistance compensation throughout the breathing cycle. By comparison, the ATC prototype ventilator compensated 79% of the effective tube resistance.

### Intrinsic PEEP, added end-inspiratory pressure, transition delay, and tidal volume in ATC

A large effective resistance, in combination with the compliance of the respiratory system, can lead to a build-up of intrinsic PEEP (iPEEP) if the expiratory time is too short for alveolar pressure equilibration. In the low- and moderate-effort conditions, the expiratory time of 2 s was sufficiently long to allow for near-complete expiration, so that added PEEP - the difference between iPEEP and set PEEP - was small (Fig. 5A, S1A). For the strong-effort pattern, however, the shorter expiratory time of 1 s, in combination with a larger tidal volume and substantial residual effective resistance, led to added PEEP in all commercial ventilators, which exceeded 6 mbar in some cases. Added PEEP was particularly pronounced when expiratory tube compensation was absent, as was the case at PEEP 0 in all commercial ventilators, and for all PEEP levels in the Evita 4 and Evita XL. With increasing PEEP, the Evita V600 provided progressively more effective expiratory tube compensation, resulting in progressively less added PEEP. During strong effort, the prototype ventilator showed an end-expiratory tracheal pressure approximately 1.2 mbar below PEEP because tracheal pressure did not return quickly enough to the set PEEP level by the end of expiration (see also Fig. 3).

**Fig. 5:**
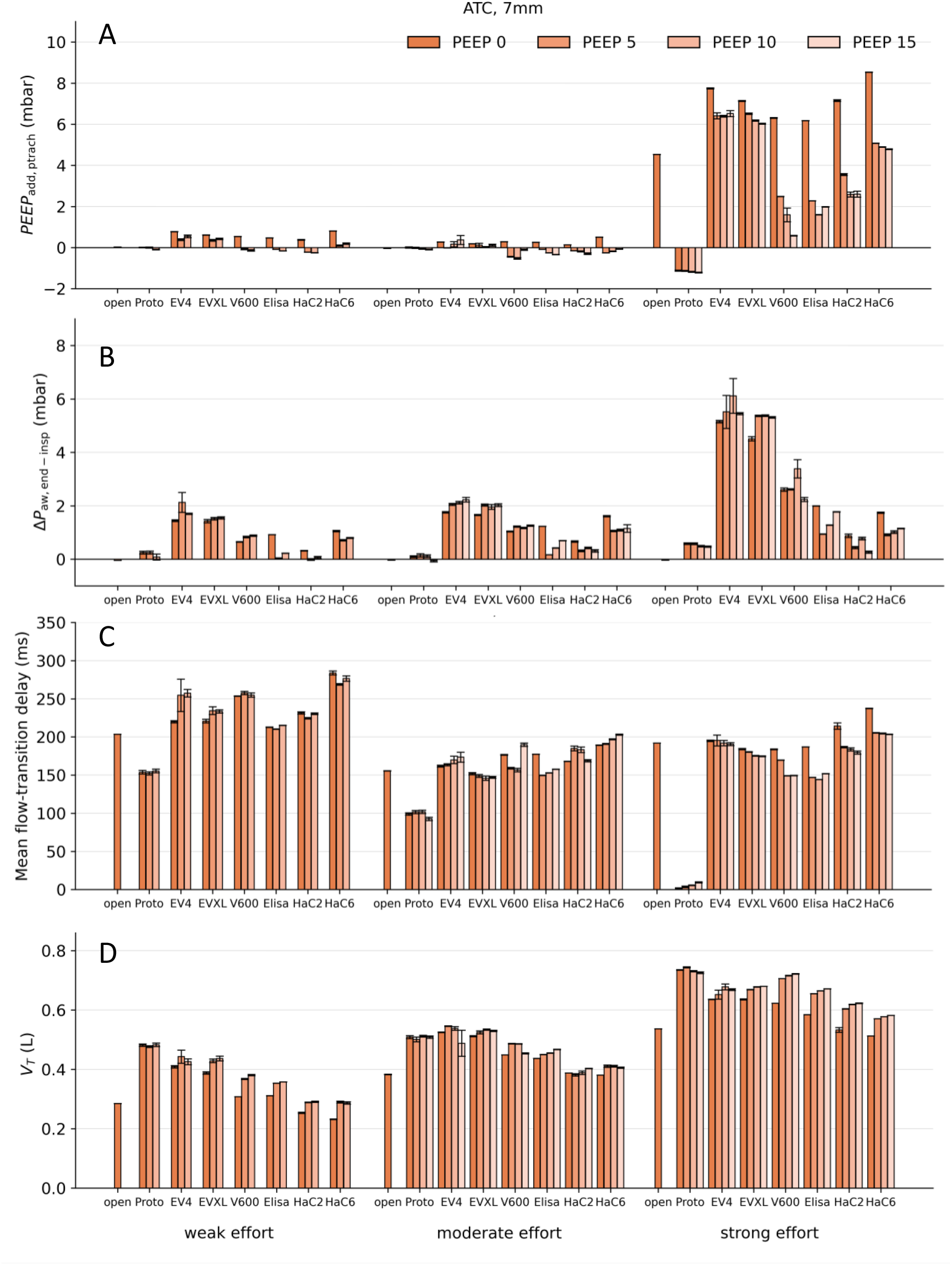
Pressure, delay, and volume parameters during ATC (7 mm ETT). (A) Tracheally measured added PEEP, computed as iPEEP - PEEP_set_; (B) end-inspiratory airway-pressure overshoot above set PEEP; (C) mean flow-transition delay, calculated as the average of the delays from the onset of inspiratory and expiratory effort to the corresponding flow transitions; and (D) tidal volume. Values are mean ± SEM from 10 ± 3 breaths per condition. The corresponding data for the 8-mm ETT are shown in Fig. S1.

The counterpart to added PEEP is an end-inspiratory airway-pressure overshoot, ΔP_aw,end-insp_, defined as end-inspiratory airway pressure above the set PEEP level. This overshoot results mainly from the finite response time of the demand-flow controller. Like added PEEP, a positive ΔP_aw,end-insp_ contributes to an increased pressure load on the lungs. We found that ΔP_aw,end-insp_ remained below approximately 0.9 mbar with Hamilton C2 and 1.8 mbar with Hamilton C6 (Fig. 5B, S1B) mainly because their tube compensation was limited and peak airway pressure rarely exceeded PEEP by more than approximately 2 mbar during inspiration (see also Fig. 3). The Evita ventilators provided better inspiratory tube compensation, with mid-inspiratory peak airway pressures rising substantially above PEEP, but this was accompanied by an increased end-inspiratory pressure overshoot, particularly during strong effort, where it reached up to 6 mbar. By contrast, the prototype ventilator kept this pressure overshoot at or below 0.6 mbar despite the even larger mid-inspiratory airway-pressure excursions required for sufficient tube compensation.

The effective resistance, together with the compliance of the respiratory system, introduces a phase shift between effort and flow. We characterized this shift by the mean transition delay, calculated as the average of the delays from the beginning and end of the inspiratory effort to the corresponding inspiratory and expiratory flow transitions. During unsupported breathing through the endotracheal tube open to atmosphere, this delay was approximately 140-200 ms (Fig. 5C, S1C). Added end-expiratory pressure can delay the inspiratory flow transition, whereas added end-inspiratory pressure can delay the expiratory flow transition. A prolonged transition delay increases the patient-ventilator asynchrony and wastes part of the patient’s inspiratory effort. With the commercial ventilators, the transition delay was roughly comparable to that during unsupported breathing. By contrast, the prototype ventilator substantially reduced the mean transition delay.

The tidal volumes for a given breathing pattern differed substantially among the tested ventilators. A higher effective resistance tends to decrease tidal volume, whereas a larger end-inspiratory airway-pressure overshoot tends to increase it. The net effect, shown in Fig. 5D, was therefore mixed: the two Hamilton ventilators, with their high effective resistance and small pressure overshoot, produced the smallest tidal volumes, comparable to those during unsupported breathing. The Dräger ventilators, with their lower effective resistance and larger end-inspiratory pressure overshoot, produced larger tidal volumes. Despite its negligible end-inspiratory pressure overshoot, the prototype ventilator achieved tidal volumes comparable to or greater than those of the Dräger ventilators.

### Effective inspiratory resistance, pressure-time-product, and added work-of-breathing

During passive expiration, the absence of expiratory tube compensation should not directly increase patient effort, except indirectly through the development of intrinsic PEEP. Thus, effective resistance calculated over the entire breath may overstate the resistive load relevant to inspiration in ventilators without expiratory tube compensation (Evita 4 and XL). We therefore also evaluated the inspiration-only effective tracheal resistance, R_eff,trach,in_, over the simulator-defined inspiratory-effort interval, with iPEEP serving as the reference pressure (see Methods). R_eff,trach,in_ is identical to the inspiratory pressure-time product (PTP) normalized to the inspired volume over the same interval.

The two older Evita 4 and XL models were the only commercial ventilators in which effective inspiratory resistance was appreciably lower than whole-breath effective resistance under both weak and moderate effort (Fig. 6, Table S1). During strong effort, effective inspiratory resistance generally exceeded whole-breath effective resistance in all commercial ventilators, with the largest increases observed in the Hamilton ventilators. By contrast, the ATC prototype maintained a low effective inspiratory resistance under all tested conditions, also during strong effort.

**Fig. 6:**
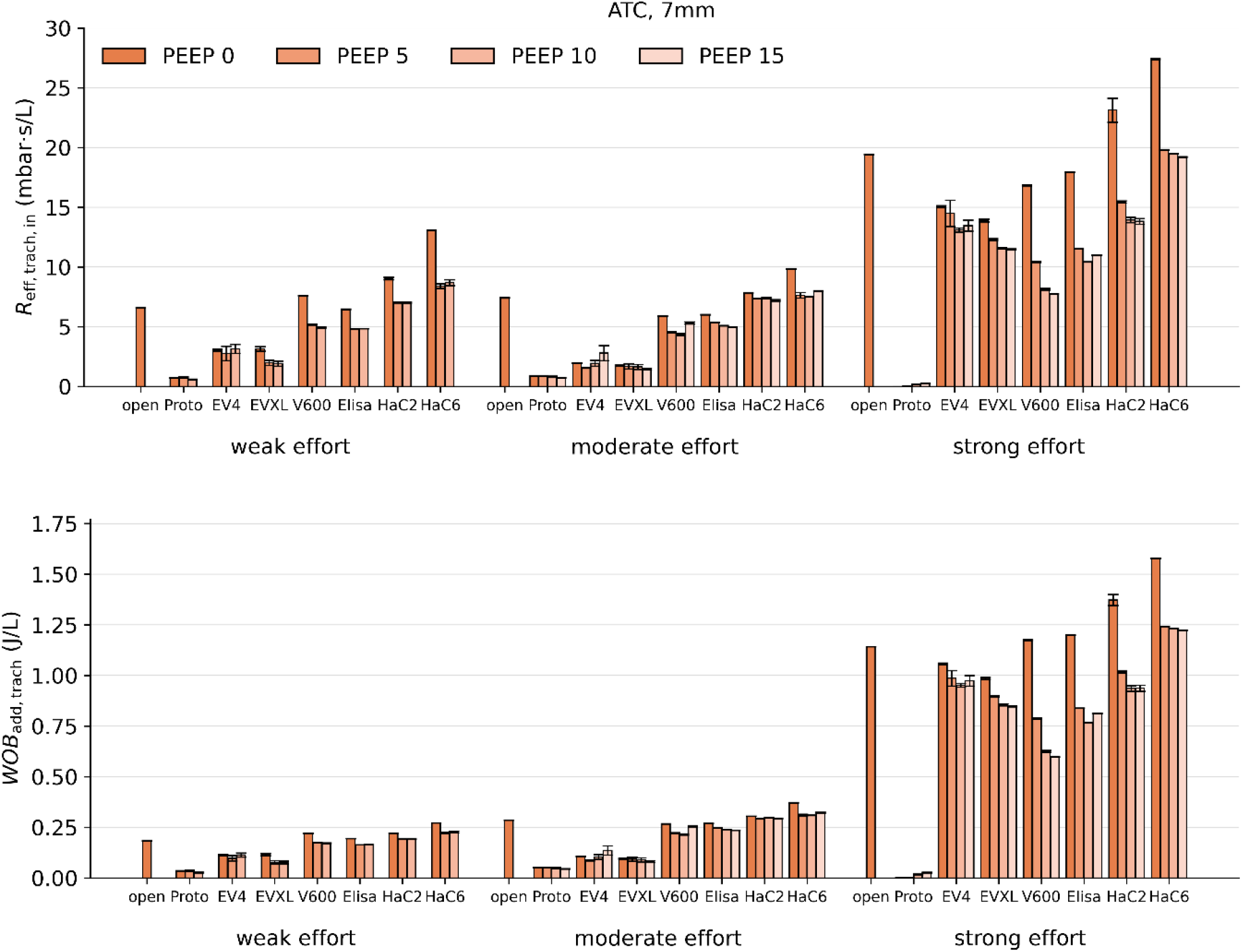
Inspiratory pressure time product and added work-of-breathing (7 mm ETT). Top: Effective tracheal resistance R_eff,trach,in_, which is the volume-normalized PTP, in inspiration during ATC with a 7 mm endotracheal tube, shown for weak, moderate, and strong inspiratory effort, different PEEP levels, and all tested ventilators. Bottom: Volume-normalized added work-of-breathing. Values are mean ± SEM from 10 ± 3 breaths per condition. The corresponding data for the 8-mm ETT are shown in Fig. S2.

Effective inspiratory resistance, or PTP, characterizes the additional inspiratory pressure burden caused by an uncompensated resistance. For comparison, we also calculated added work of breathing (WOB_add_), which quantifies the wasted mechanical work per unit inspired volume required to overcome the uncompensated resistance of the endotracheal tube-ventilator system. As expected, WOB_add_ closely paralleled the PTP within each effort condition but increased more steeply with effort intensity because it additionally depends on inspiratory flow (Figs. 6 and S2).

## Discussion

Our data show that despite a nominal 100% ATC setting, the tested commercial ventilators left most of the resistive load imposed by the endotracheal tube uncompensated. Even with PEEP 0 excluded, their average reduction in whole-breath effective resistance relative to T-piece breathing did not exceed 27%, and some models even increased the load.

A substantial part of the incomplete tube compensation appears to result from limitations in airway pressure control. In CPAP mode, all tested commercial ventilators showed marked deviations of airway pressure from the target pressure. These deviations were particularly large in the two Hamilton ventilators and corresponded to an effective resistance from the ventilator alone of more than 3 mbar·s/L.

In the Dräger and Löwenstein ventilators, the total effective resistance for a 7 mm tube under ATC reached, on average, more than 4 mbar·s/L at weak inspiratory effort, approximately 6 mbar·s/L at moderate effort, and approximately 8 mbar·s/L at strong effort. In the two Hamilton ventilators, the effective resistance was about 2 mbar·s/L higher still, exceeding 10 mbar·s/L at high respiratory demand. Thus, the highest residual loads occurred under the condition in which mechanical unloading matters most for the patient. Taking the resistance of an N95 facemask (approximately 0.5 mbar·s/L [13]) as a reference, the residual load in ATC mode would roughly correspond to the resistance encountered when breathing through 12-20 facemasks stacked together.

These results broadly confirm an earlier bench study from 2003 that compared commercial ventilators with an ATC prototype ventilator [11]. However, our results seem to conflict with a more recent bench comparison of seven ICU ventilators [14]. This study found only small differences in tidal volume between breathing with 100% ATC through an 8-mm endotracheal tube versus a no-tube reference condition. However, tidal volume was the study’s primary endpoint under a single patient-derived effort sequence and one PEEP level. As our data demonstrate, preserved tidal volume does not establish effective tube compensation, because residual resistance, transition delay, intrinsic PEEP, and airway-pressure overshoot can have opposing effects on volume.

The reasons for the limited tracheal-pressure control observed in the commercial ventilators remain unclear. The performance of the ATC prototype ventilator demonstrates that effective tracheal-pressure control is technically feasible. In addition, the two older-generation ventilators, Evita 4 and Evita XL, showed reasonable tube compensation during inspiration. This raises the question why, despite satisfactory inspiratory tube compensation, the inspiratory pressure-time-product and added work-of-breathing were so high in both of these commercial ventilators during strong efforts. Our results point to the absence of expiratory tube compensation and the resulting rise in intrinsic PEEP as the main reason.

Full expiratory tube compensation requires the ventilator to lower airway pressure below PEEP and, at low PEEP, sometimes below atmospheric pressure. Both the original ATC prototype and the current ATC prototype were able to do so. This capability requires additional safety precautions, because valve failure could expose the patient to harmful negative airway pressures. It is therefore understandable that manufacturers have avoided active subatmospheric pressure control. However, all four contemporary ventilators tested here offered expiratory tube compensation at higher PEEP levels, where subatmospheric pressure was not required. Even under these conditions, only the Evita V600 and Elisa 800 lowered expiratory airway pressure appreciably below PEEP.

Our study introduces the effective resistance as a measure of how accurately ventilators control airway pressure in CPAP mode and tracheal pressure in ATC mode. To our knowledge, the effective resistance has not previously been used for this purpose, and directly comparable published values are therefore not available. Nonetheless, we propose its wider use as a simple and informative parameter that penalizes all deviations from the target pressure, regardless of when they occur. It can therefore be calculated without identifying the timing of inspiratory and expiratory efforts. Moreover, by normalizing pressure variability by flow variability, the effective resistance is considerably less dependent on the breathing pattern and inspiratory effort than added work of breathing (WOB_add_) and pressure-time product (PTP). Therefore, the effective resistance summarizes overall dynamic pressure-control performance but may at first appear less suited than WOB_add_ and PTP for quantifying the physiological burden imposed by the tube and the ventilator during inspiration. However, this overlooks the carry-over effects of expiratory resistance.

Our study establishes that intrinsic PEEP can substantially rise during high-intensity spontaneous breathing, and that effective expiratory tube compensation can prevent this rise. Limiting iPEEP also helps to reduce the delay in inspiratory flow delivery. Together, residual expiratory resistance, expiratory transition delay, and iPEEP contributed to the elevated PTP and WOB_add_ seen in all commercial ventilators during strong effort, at levels that are likely to cause patient discomfort and respiratory muscle fatigue. By contrast, the ATC prototype largely avoided the added resistance imposed by the tube and ventilator circuit, which reduced the transition delay and iPEEP build-up and minimized PTP and WOB_add._

Early clinical studies in intubated patients established that effective ATC can reduce tube-related work of breathing under high ventilatory demand [5, 9]. Later clinical studies of commercial ATC implementations have mainly examined its role during spontaneous breathing trials, with mixed results: several studies found no clear reduction in weaning duration or mechanical ventilation time compared with CPAP or low pressure support [15–17], whereas a meta-analysis suggested that ATC may improve extubation success compared with other spontaneous breathing trial modes [18]. None of these studies, however, investigated whether and to what extent commercial ventilators actually compensated endotracheal tube resistance. Consequently, the benefits demonstrated in the earlier patient studies with a prototype that achieved near-complete tube compensation cannot be assumed to apply to commercial implementations without evidence that they achieve comparable tracheal-pressure control. Given that ATC is a commonly used mode in clinical practice [14], a thorough clinical evaluation of effective tracheal-pressure control as provided for example with our ATC prototype is urgently needed to establish its benefits, or lack thereof, for patients, including its effects on weaning duration, breathing pattern and respiratory control, need for sedation, or risk of patient self-inflicted lung injury (PSILI).

In our bench study, we did not test the effects of breath-to-breath variability, intratidal changes in compliance, or adaptation of respiratory drive to the level of support. These factors may affect ventilator control performance, particularly if control algorithms adapt to respiratory mechanics and effort, as indicated by the improvement in Evita V600 pressure control during the first minute of simulated breathing (see Methods).

We noticed that tube compensation improved when the ventilators were set to a smaller than the actual tube diameter. This observation suggests that tube compensation may have been intentionally incomplete, possibly to avoid overcompensation or unstable pressure control. In clinical practice, however, endotracheal tube resistance is often higher than that of a clean tube because of mucus deposition, biofilm formation, or kinking [19, 20]. Added components such as an HME filter increase the total resistance further. We previously demonstrated that the combined resistance of such added components translates into a reduced effective tube diameter and hence can be compensated in ATC mode [4, 21]. However, we discourage clinicians from selecting a smaller than the nominal tube diameter unless the added resistance has been reliably measured and converted into an effective tube diameter. A method to estimate this effective diameter *in situ* has recently been published [21].

Since we tested only two tube sizes, three distinct breathing patterns, four contemporary and two older-generation commercial ventilators from three manufacturers, and used only a single geometry of the artificial trachea, our results cannot be generalized to all clinical situations and all ICU ventilators currently on the market. Nonetheless, our findings show that a nominal 100% ATC setting does not guarantee effective tube compensation. Therefore, we recommend that regulatory authorities require every ICU ventilator model to undergo testing by independent, accredited laboratories, using standardized protocols to assess pressure-control performance during spontaneous breathing in CPAP and ATC modes. The test results should be publicly accessible.

## Declarations

### Author approval

All authors have seen and approved the manuscript.

### Ethics approval and consent to participate

This study did not involve human participants, data, or identifiable images.

### Consent for publication

Not applicable. This study did not involve human participants, data, or identifiable images.

### Availability of data and materials

The raw data and analysis software can be obtained from the corresponding author upon request.

### Author Contributions

BF designed the study, CK designed and built the active lung simulator, BF and CK designed and built the prototype ventilator, BF performed the experiments and analyzed the data, BF and RF wrote the manuscript.

## Acknowledgments

We thank Christoph Gründler, Alexander Rentrop, Christoph Laußer and Thomas Flohr for providing ventilators for testing, technical support, and for helpful discussions.

## Supplementary information

**Fig. S1:**
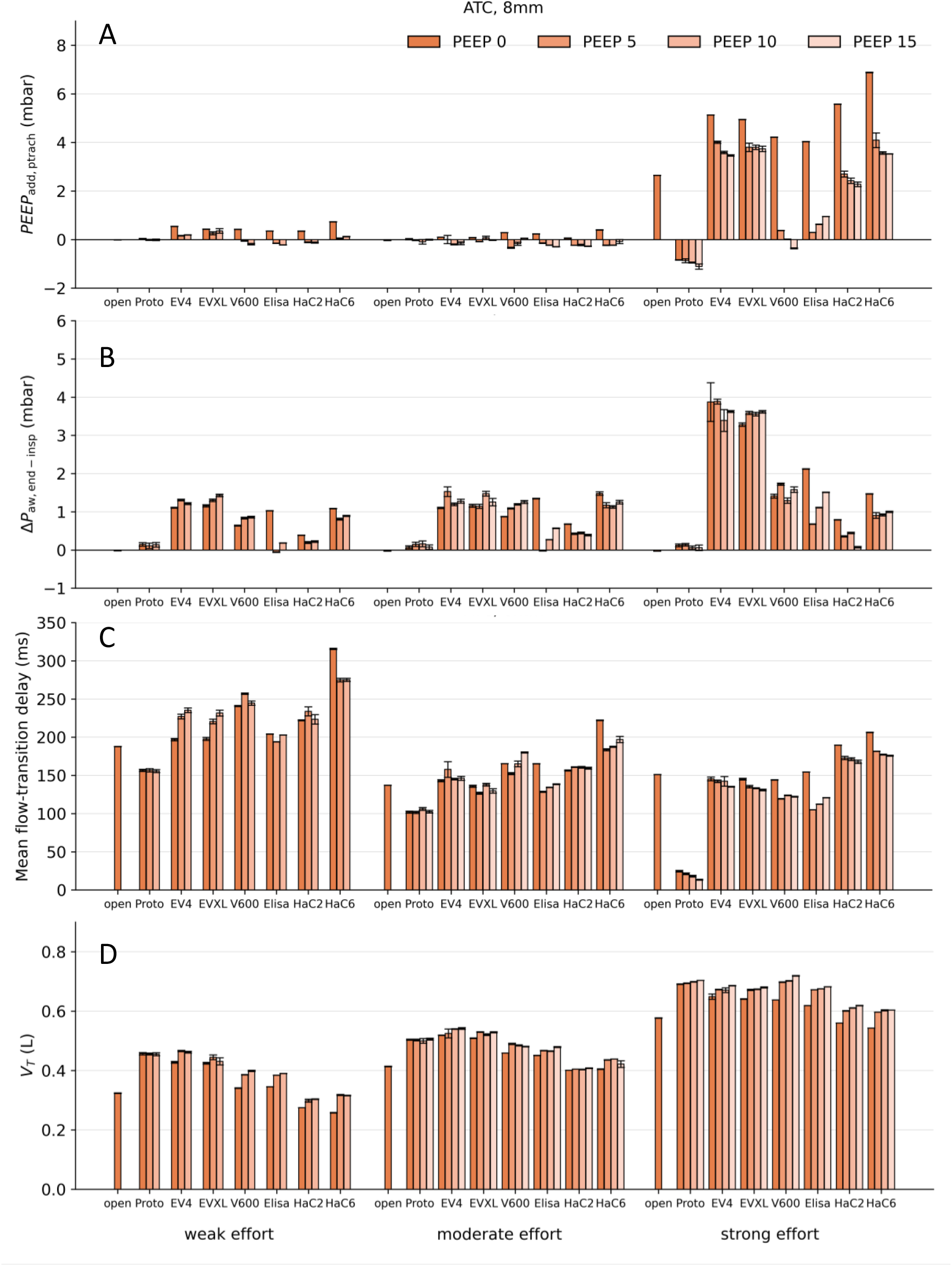
Pressure, delay, and volume parameters during ATC (8 mm ETT). (A) Tracheally measured added PEEP, computed as iPEEP - PEEP_set_; (B) end-inspiratory airway-pressure overshoot above set PEEP; (C) mean flow-transition delay, calculated as the average of the delays from the onset of inspiratory and expiratory effort to the corresponding flow transitions; and (D) tidal volume. Values are mean ± SEM from 10 ± 3 breaths per condition.

**Fig. S2:**
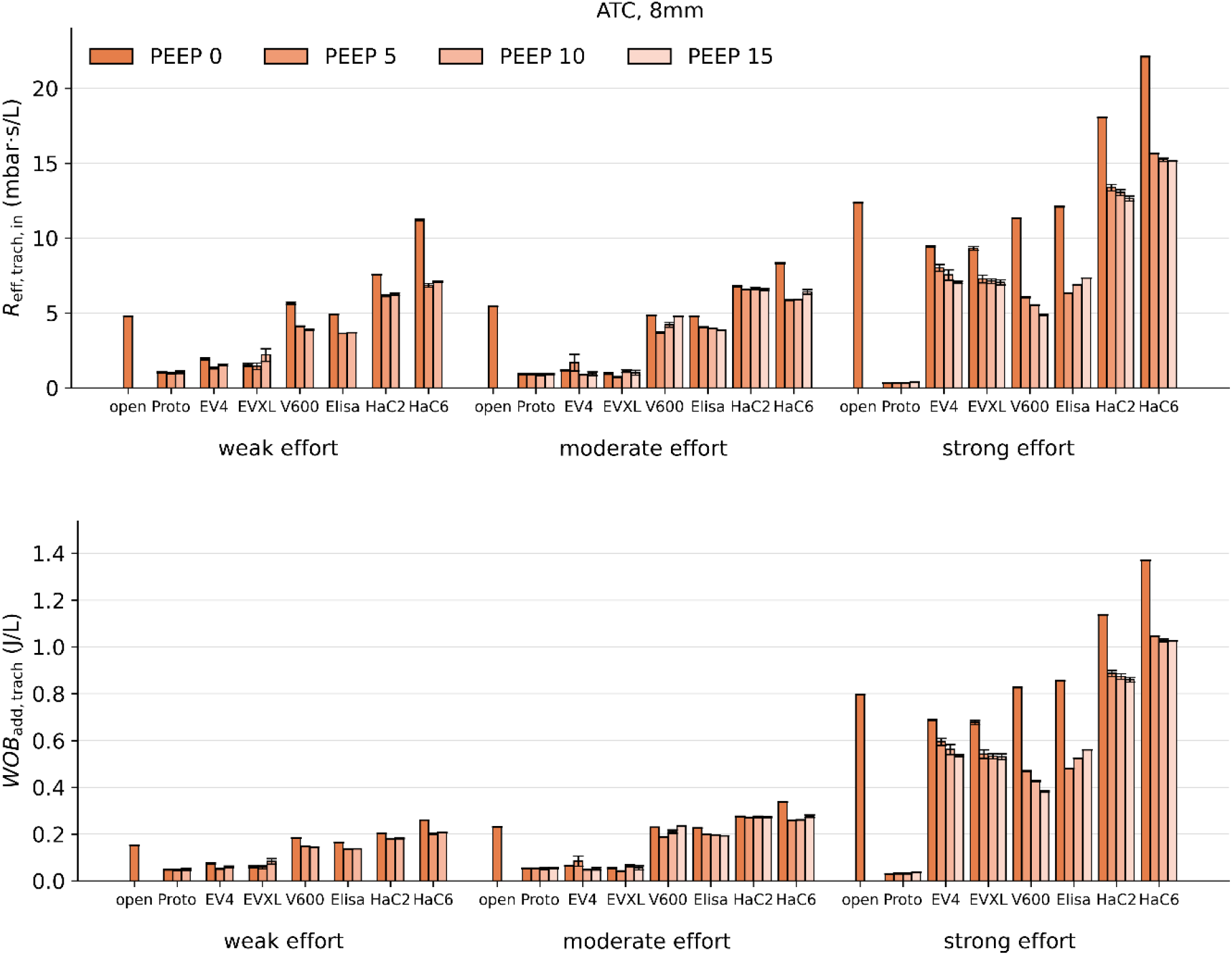
Inspiratory pressure time product and added work-of-breathing (8 mm ETT). Top: Effective tracheal resistance R_eff,trach,in_ in inspiration during ATC with an 8 mm endotracheal tube, shown for weak, moderate, and strong inspiratory effort, different PEEP levels, and all tested ventilators. R_eff,trach,in_ equals the volume-normalized inspiratory pressure-time product. Bottom: Volume-normalized added work-of-breathing. Values are mean ± SEM from 10 ± 3 breaths per condition.

**Fig. S3:**
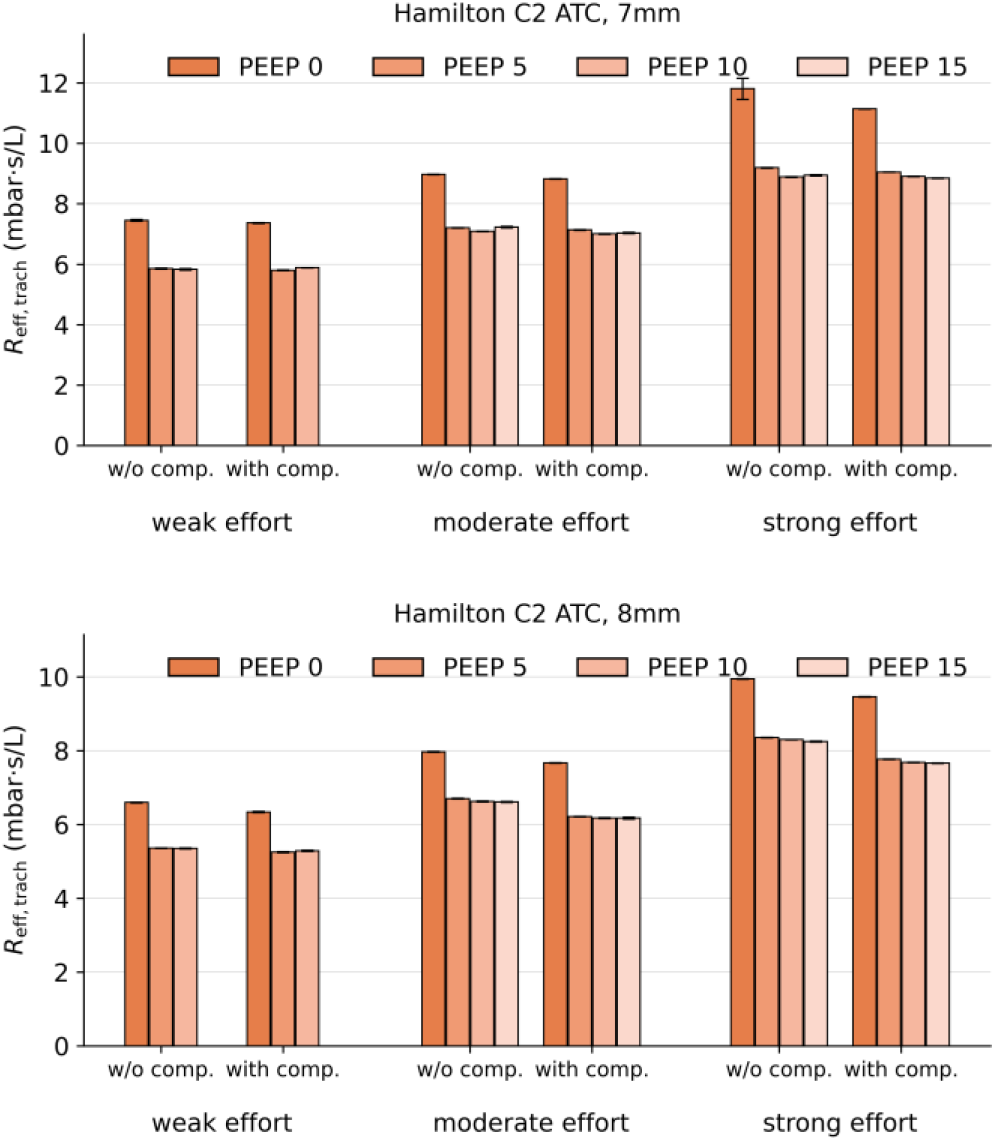
Effect of flow sensor placement on effective tracheal resistance. Both Hamilton ventilators require a variable-orifice flow sensor placed between the flexible tube and the Y-piece. This sensor has a resistance of approximately 1 mbar·s/L. The placement experiment shown here was performed with the Hamilton C2. In the standard configuration (“w/o comp.”, left bars), the Hamilton sensor was placed between the Y-piece and our thermal mass flow sensor and airway-pressure adapter. This made the measurements comparable across ventilators and excluded the pressure drop across the Hamilton sensor from our airway and tracheal pressure measurements, but left the small resistance of our own measuring assembly uncompensated. In the alternative configuration (“with comp.”, right bars), the Hamilton sensor was placed between our measuring assembly and the flexible tube, which allowed the ventilator’s demand-flow controller to compensate the resistance of our assembly. R_eff,trach_ was largely independent of sensor placement except during strong effort when compensation reduced R_eff,trach_ by up to 0.5 mbar·s/L.

**Fig. S4:**
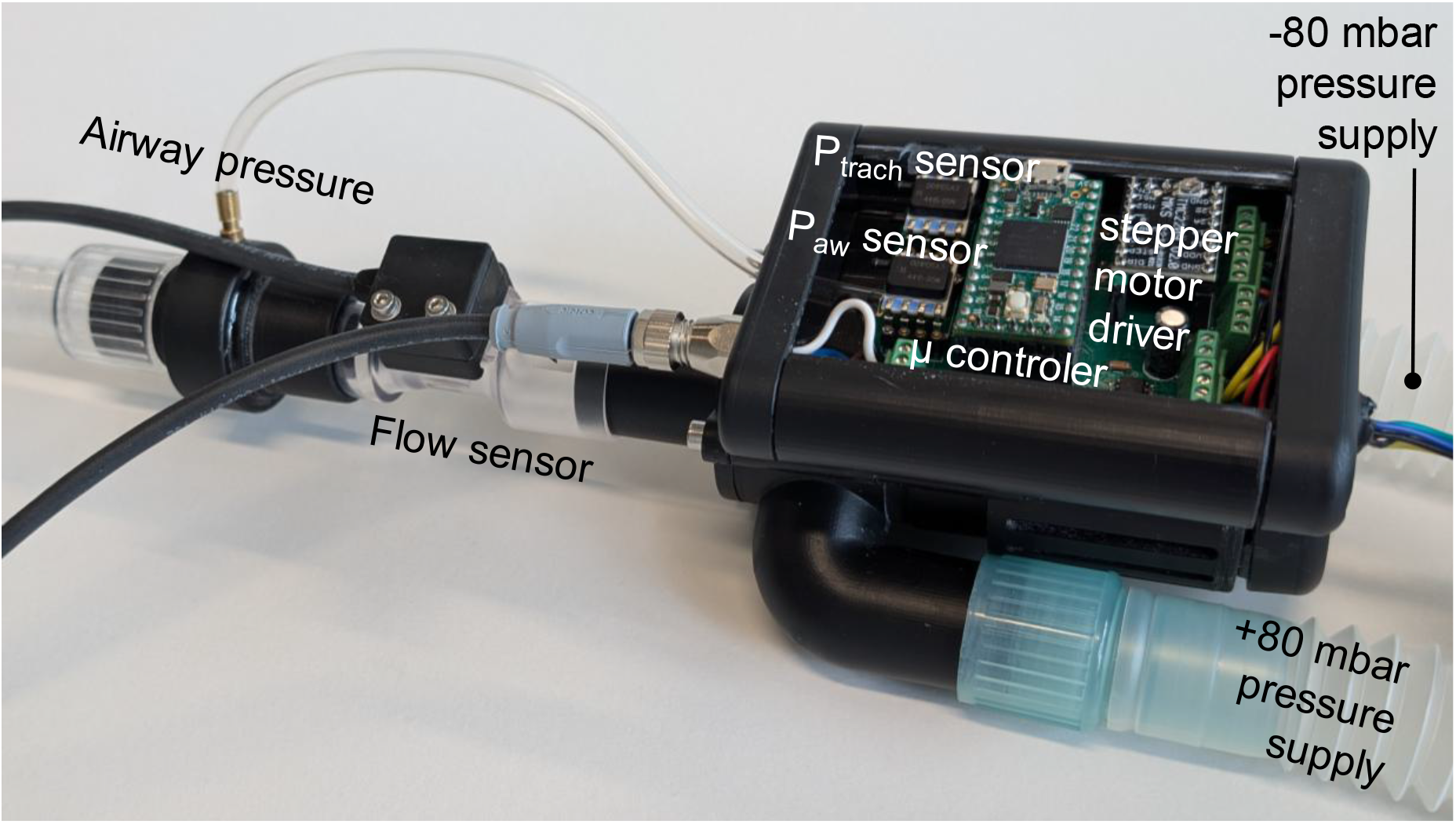
Photograph of the ATC prototype ventilator. The prototype ventilator replaces the traditional Y-piece with a rotary valve driven by a stepper motor. Inspiratory and expiratory pressure of +80 and −80 mbar, respectively, is supplied by radial blowers (not shown). Flow and airway pressure sensors are placed in close proximity to the ventilator to improve response time.

**Table S1:** Characterization of intrinsic PEEP, effective resistance, effective resistance during inspiration (= volume-normalized pressure-time-product), added work of breathing, and tidal volume for different tube sizes, breathing efforts, ventilators, and modes.

| ETT | Effort | Ventilator | Mode | PEEP | iPEEP | ΔP<sub>aw, end-insp</sub> | R<sub>eff,aw</sub> | R<sub>eff,trach</sub> | R<sub>eff,trach,in</sub><br>(= PTP) | WOB<sub>add</sub> | V<sub>T</sub> | flow delay |
|---|---|---|---|---|---|---|---|---|---|---|---|---|
| *mm* |  |  |  | *mbar* | *mbar* | *mbar* | *mbar·s/L* | *mbar·s/L* | *mbar·s/L* | *J/L* | *mL* | *ms* |
| **7** | **weak** | **TPiece** | **TPiece** | 0 | 0.0 |  |  | 5.87 | 6.61 | 0.18 | 285 | 204 |
|  |  | **Prototype** | **CPAP** | 0 | 0.1 | 0.1 | 0.46 | 5.59 | 6.26 | 0.18 | 300 | 205 |
|  |  |  |  | 5 | 5.1 | 0.2 | 0.44 | 5.60 | 6.20 | 0.18 | 305 | 205 |
|  |  |  |  | 10 | 10.1 | 0.3 | 0.45 | 5.61 | 6.30 | 0.19 | 306 | 207 |
|  |  |  | **ATC** | 0 | 0.0 | 0.2 |  | 1.28 | 0.74 | 0.04 | 482 | 154 |
|  |  |  |  | 5 | 5.0 | 0.3 |  | 1.30 | 0.76 | 0.04 | 477 | 152 |
|  |  |  |  | 10 | 9.9 | 0.1 |  | 1.31 | 0.58 | 0.03 | 483 | 155 |
|  |  | **EVITA4** | **CPAP** | 0 | 0.4 | 0.8 | 2.32 | 7.46 | 9.27 | 0.23 | 260 | 270 |
|  |  |  |  | 5 | 5.2 | 0.8 | 1.93 | 7.02 | 8.91 | 0.22 | 268 | 282 |
|  |  |  |  | 10 | 10.3 | 0.7 | 2.06 | 7.16 | 9.34 | 0.23 | 266 | 286 |
|  |  |  | **ATC** | 0 | 0.8 | 1.4 |  | 4.28 | 3.03 | 0.11 | 409 | 220 |
|  |  |  |  | 5 | 5.4 | 2.1 |  | 4.37 | 2.77 | 0.10 | 443 | 255 |
|  |  |  |  | 10 | 10.6 | 1.7 |  | 4.22 | 3.15 | 0.11 | 426 | 257 |
|  |  | **EVITAXL** | **CPAP** | 0 | 0.4 | 0.8 | 2.18 | 7.34 | 9.06 | 0.22 | 262 | 268 |
|  |  |  |  | 5 | 5.4 | 0.9 | 1.59 | 6.72 | 8.97 | 0.22 | 267 | 273 |
|  |  |  |  | 10 | 10.2 | 0.9 | 1.42 | 6.58 | 7.84 | 0.21 | 285 | 265 |
|  |  |  | **ATC** | 0 | 0.6 | 1.4 |  | 4.45 | 3.15 | 0.12 | 388 | 221 |
|  |  |  |  | 5 | 5.4 | 1.5 |  | 3.89 | 1.99 | 0.08 | 429 | 235 |
|  |  |  |  | 10 | 10.4 | 1.5 |  | 3.73 | 1.93 | 0.08 | 437 | 234 |
|  |  | **Evita V600** | **CPAP** | 0 | 0.5 | 0.7 | 1.88 | 6.84 | 9.24 | 0.24 | 270 | 262 |
|  |  |  |  | 5 | 5.1 | 0.6 | 1.74 | 6.58 | 8.09 | 0.22 | 288 | 267 |
|  |  |  |  | 10 | 10.1 | 0.6 | 1.68 | 6.48 | 8.09 | 0.22 | 293 | 261 |
|  |  |  | **ATC** | 0 | 0.5 | 0.7 |  | 5.95 | 7.61 | 0.22 | 308 | 254 |
|  |  |  |  | 5 | 4.9 | 0.8 |  | 4.61 | 5.20 | 0.18 | 368 | 258 |
|  |  |  |  | 10 | 9.9 | 0.9 |  | 4.54 | 4.93 | 0.17 | 381 | 255 |
|  |  | **Elisa 800** | **CPAP** | 0 | 0.5 | 0.8 | 2.01 | 6.97 | 8.79 | 0.22 | 270 | 228 |
|  |  |  |  | 5 | 5.1 | -0.1 | 0.68 | 5.88 | 7.16 | 0.20 | 293 | 207 |
|  |  |  |  | 10 | 10.0 | 0.1 | 0.92 | 6.06 | 7.05 | 0.20 | 296 | 221 |
|  |  |  | **ATC** | 0 | 0.5 | 0.9 |  | 5.88 | 6.47 | 0.20 | 311 | 213 |
|  |  |  |  | 5 | 4.9 | 0.0 |  | 4.30 | 4.82 | 0.16 | 354 | 210 |
|  |  |  |  | 10 | 9.8 | 0.2 |  | 4.49 | 4.85 | 0.17 | 358 | 215 |
|  |  | **Hamilton C2** | **CPAP** | 0 | 0.3 | 0.2 | 3.42 | 8.33 | 10.26 | 0.23 | 233 | 232 |
|  |  |  |  | 5 | 4.8 | 0.0 | 2.54 | 7.66 | 9.35 | 0.22 | 246 | 237 |
|  |  |  |  | 10 | 9.8 | 0.0 | 2.70 | 7.72 | 9.45 | 0.22 | 245 | 239 |
|  |  |  | **ATC** | 0 | 0.4 | 0.3 |  | 7.46 | 9.05 | 0.22 | 254 | 232 |
|  |  |  |  | 5 | 4.8 | 0.0 |  | 5.86 | 7.03 | 0.19 | 289 | 225 |
|  |  |  |  | 10 | 9.8 | 0.1 |  | 5.83 | 7.00 | 0.19 | 291 | 231 |
|  |  | **Hamilton C6** | **CPAP** | 0 | 0.7 | 0.9 | 6.50 | 10.96 | 16.73 | 0.30 | 198 | 330 |
|  |  |  |  | 5 | 5.1 | 0.6 | 3.48 | 8.35 | 10.95 | 0.25 | 250 | 291 |
|  |  |  |  | 10 | 10.2 | 0.7 | 3.57 | 8.36 | 11.18 | 0.25 | 250 | 302 |
|  |  |  | **ATC** | 0 | 0.8 | 1.1 |  | 9.09 | 13.07 | 0.27 | 232 | 284 |
|  |  |  |  | 5 | 5.1 | 0.7 |  | 6.79 | 8.41 | 0.22 | 290 | 269 |
|  |  |  |  | 10 | 10.2 | 0.8 |  | 6.88 | 8.70 | 0.23 | 287 | 277 |
| **7** | **moderate** | **TPiece** | **TPiece** | 0 | 0.0 |  |  | 7.64 | 7.43 | 0.29 | 383 | 156 |
|  |  | **Prototype** | **CPAP** | 0 | 0.0 | 0.3 | 0.52 | 7.38 | 7.30 | 0.29 | 399 | 159 |
|  |  |  |  | 5 | 5.0 | 0.3 | 0.54 | 7.40 | 7.24 | 0.29 | 396 | 160 |
|  |  |  |  | 10 | 10.0 | 0.3 | 0.62 | 7.38 | 7.26 | 0.29 | 395 | 159 |
|  |  |  |  | 15 | 15.0 | 0.4 | 0.54 | 7.35 | 7.20 | 0.29 | 398 | 160 |
|  |  |  | **ATC** | 0 | 0.0 | 0.1 |  | 1.69 | 0.89 | 0.05 | 509 | 99 |
|  |  |  |  | 5 | 5.0 | 0.1 |  | 1.71 | 0.87 | 0.05 | 501 | 102 |
|  |  |  |  | 10 | 10.0 | 0.1 |  | 1.71 | 0.83 | 0.05 | 512 | 102 |
|  |  |  |  | 15 | 14.9 | -0.1 |  | 1.67 | 0.74 | 0.05 | 509 | 93 |
|  |  | **EVITA4** | **CPAP** | 0 | 0.1 | 1.1 | 2.57 | 9.38 | 8.95 | 0.34 | 377 | 202 |
|  |  |  |  | 5 | 4.9 | 1.1 | 1.85 | 8.78 | 8.45 | 0.32 | 386 | 201 |
|  |  |  |  | 10 | 9.9 | 1.2 | 1.85 | 8.76 | 8.49 | 0.33 | 388 | 200 |
|  |  |  |  | 15 | 15.0 | 1.2 | 1.97 | 8.83 | 8.61 | 0.33 | 387 | 203 |
|  |  |  | **ATC** | 0 | 0.3 | 1.8 |  | 6.10 | 1.96 | 0.11 | 525 | 162 |
|  |  |  |  | 5 | 5.0 | 2.1 |  | 5.73 | 1.58 | 0.09 | 546 | 164 |
|  |  |  |  | 10 | 10.2 | 2.1 |  | 5.75 | 1.94 | 0.11 | 538 | 170 |
|  |  |  |  | 15 | 15.4 | 2.2 |  | 6.16 | 2.80 | 0.14 | 488 | 174 |
|  |  | **EVITAXL** | **CPAP** | 0 | 0.2 | 1.1 | 2.40 | 9.24 | 8.83 | 0.33 | 378 | 197 |
|  |  |  |  | 5 | 5.0 | 1.1 | 1.53 | 8.48 | 8.41 | 0.32 | 386 | 196 |
|  |  |  |  | 10 | 10.0 | 1.2 | 1.46 | 8.40 | 8.45 | 0.32 | 387 | 198 |
|  |  |  |  | 15 | 15.1 | 1.3 | 1.49 | 8.40 | 8.43 | 0.32 | 389 | 196 |
|  |  |  | **ATC** | 0 | 0.2 | 1.7 |  | 5.99 | 1.76 | 0.10 | 512 | 152 |
|  |  |  |  | 5 | 5.1 | 2.0 |  | 5.71 | 1.69 | 0.09 | 525 | 149 |
|  |  |  |  | 10 | 10.0 | 2.0 |  | 5.60 | 1.65 | 0.09 | 534 | 146 |
|  |  |  |  | 15 | 15.1 | 2.0 |  | 5.51 | 1.46 | 0.08 | 530 | 147 |
|  |  | **Evita V600** | **CPAP** | 0 | 0.3 | 0.8 | 1.57 | 8.41 | 8.53 | 0.33 | 393 | 195 |
|  |  |  |  | 5 | 4.9 | 0.9 | 1.40 | 7.99 | 8.10 | 0.32 | 401 | 193 |
|  |  |  |  | 10 | 9.7 | 0.9 | 1.67 | 7.93 | 7.72 | 0.31 | 409 | 181 |
|  |  |  |  | 15 | 14.5 | 0.8 | 2.06 | 7.98 | 7.78 | 0.32 | 413 | 179 |
|  |  |  | **ATC** | 0 | 0.3 | 1.0 |  | 7.26 | 5.90 | 0.27 | 449 | 177 |
|  |  |  |  | 5 | 4.6 | 1.2 |  | 5.87 | 4.53 | 0.22 | 487 | 159 |
|  |  |  |  | 10 | 9.5 | 1.2 |  | 5.80 | 4.36 | 0.22 | 486 | 157 |
|  |  |  |  | 15 | 14.9 | 1.3 |  | 5.76 | 5.32 | 0.26 | 454 | 190 |
|  |  | **Elisa 800** | **CPAP** | 0 | 0.3 | 1.2 | 2.05 | 8.63 | 8.64 | 0.34 | 394 | 194 |
|  |  |  |  | 5 | 5.1 | -0.1 | 0.91 | 7.65 | 8.11 | 0.32 | 386 | 172 |
|  |  |  |  | 10 | 9.8 | 0.2 | 1.21 | 7.87 | 7.70 | 0.31 | 405 | 170 |
|  |  |  |  | 15 | 14.8 | 0.4 | 1.40 | 7.98 | 7.57 | 0.31 | 411 | 172 |
|  |  |  | **ATC** | 0 | 0.3 | 1.2 |  | 7.48 | 6.02 | 0.27 | 437 | 177 |
|  |  |  |  | 5 | 4.9 | 0.2 |  | 5.54 | 5.35 | 0.25 | 450 | 150 |
|  |  |  |  | 10 | 9.8 | 0.4 |  | 5.75 | 5.09 | 0.24 | 455 | 153 |
|  |  |  |  | 15 | 14.7 | 0.7 |  | 5.98 | 4.97 | 0.24 | 467 | 158 |
|  |  | **Hamilton C2** | **CPAP** | 0 | 0.1 | 0.3 | 3.68 | 10.23 | 10.17 | 0.35 | 345 | 174 |
|  |  |  |  | 5 | 4.7 | 0.1 | 3.09 | 9.75 | 10.15 | 0.35 | 325 | 194 |
|  |  |  |  | 10 | 9.8 | 0.2 | 2.92 | 9.57 | 10.41 | 0.36 | 300 | 205 |
|  |  |  |  | 15 | 14.8 | 0.2 | 2.88 | 9.53 | 10.42 | 0.36 | 301 | 206 |
|  |  |  | **ATC** | 0 | 0.1 | 0.7 |  | 8.97 | 7.84 | 0.31 | 388 | 168 |
|  |  |  |  | 5 | 4.8 | 0.3 |  | 7.20 | 7.36 | 0.29 | 382 | 185 |
|  |  |  |  | 10 | 9.8 | 0.4 |  | 7.09 | 7.40 | 0.30 | 389 | 183 |
|  |  |  |  | 15 | 14.7 | 0.3 |  | 7.23 | 7.22 | 0.29 | 403 | 169 |
|  |  | **Hamilton C6** | **CPAP** | 0 | 0.4 | 1.4 | 6.25 | 12.37 | 14.06 | 0.44 | 324 | 248 |
|  |  |  |  | 5 | 4.8 | 1.1 | 3.41 | 9.98 | 10.23 | 0.37 | 372 | 218 |
|  |  |  |  | 10 | 9.8 | 1.2 | 3.26 | 9.87 | 9.97 | 0.37 | 371 | 216 |
|  |  |  |  | 15 | 15.0 | 1.4 | 3.27 | 9.86 | 10.14 | 0.37 | 370 | 218 |
|  |  |  | **ATC** | 0 | 0.5 | 1.6 |  | 10.27 | 9.86 | 0.37 | 381 | 189 |
|  |  |  |  | 5 | 4.8 | 1.1 |  | 8.34 | 7.65 | 0.31 | 411 | 191 |
|  |  |  |  | 10 | 9.8 | 1.1 |  | 8.04 | 7.54 | 0.31 | 411 | 197 |
|  |  |  |  | 15 | 14.9 | 1.2 |  | 8.10 | 7.99 | 0.32 | 406 | 203 |
| **7** | **strong** | **TPiece** | **TPiece** | 0 | 4.5 |  |  | 10.67 | 19.43 | 1.14 | 537 | 192 |
|  |  | **Prototype** | **CPAP** | 0 | 4.6 | 0.4 | 0.46 | 10.38 | 18.74 | 1.18 | 579 | 199 |
|  |  |  |  | 5 | 9.5 | 0.4 | 0.47 | 10.38 | 18.51 | 1.18 | 585 | 195 |
|  |  |  |  | 10 | 14.6 | 0.4 | 0.46 | 10.41 | 18.55 | 1.19 | 591 | 193 |
|  |  |  |  | 15 | 19.5 | 0.4 | 0.47 | 10.42 | 18.35 | 1.19 | 597 | 193 |
|  |  |  | **ATC** | 0 | -1.1 | 0.6 |  | 1.25 | 0.01 | 0.00 | 735 | 1 |
|  |  |  |  | 5 | 3.9 | 0.6 |  | 1.32 | 0.03 | 0.00 | 744 | 4 |
|  |  |  |  | 10 | 8.8 | 0.5 |  | 1.47 | 0.18 | 0.02 | 731 | 6 |
|  |  |  |  | 15 | 13.8 | 0.5 |  | 1.53 | 0.27 | 0.03 | 726 | 10 |
|  |  | **EVITA4** | **CPAP** | 0 | 6.9 | 1.4 | 2.85 | 12.78 | 25.90 | 1.46 | 516 | 236 |
|  |  |  |  | 5 | 10.7 | 1.9 | 2.34 | 12.17 | 23.07 | 1.35 | 547 | 231 |
|  |  |  |  | 10 | 15.8 | 1.5 | 2.02 | 12.05 | 23.74 | 1.38 | 541 | 230 |
|  |  |  |  | 15 | 20.4 | 1.6 | 1.98 | 12.00 | 22.80 | 1.35 | 549 | 228 |
|  |  |  | **ATC** | 0 | 7.7 | 5.2 |  | 8.72 | 15.06 | 1.06 | 636 | 195 |
|  |  |  |  | 5 | 11.4 | 5.5 |  | 8.91 | 14.50 | 0.99 | 652 | 195 |
|  |  |  |  | 10 | 16.4 | 6.1 |  | 8.44 | 13.10 | 0.95 | 678 | 192 |
|  |  |  |  | 15 | 21.5 | 5.5 |  | 8.23 | 13.47 | 0.97 | 669 | 191 |
|  |  | **EVITAXL** | **CPAP** | 0 | 6.3 | 1.2 | 2.55 | 12.45 | 24.48 | 1.40 | 526 | 225 |
|  |  |  |  | 5 | 10.3 | 1.5 | 1.99 | 11.94 | 22.19 | 1.31 | 551 | 220 |
|  |  |  |  | 10 | 15.2 | 1.5 | 1.87 | 11.84 | 21.77 | 1.30 | 554 | 216 |
|  |  |  |  | 15 | 20.4 | 1.5 | 1.79 | 11.75 | 22.13 | 1.33 | 556 | 217 |
|  |  |  | **ATC** | 0 | 7.1 | 4.5 |  | 8.33 | 13.89 | 0.99 | 636 | 184 |
|  |  |  |  | 5 | 11.5 | 5.4 |  | 7.89 | 12.31 | 0.90 | 669 | 181 |
|  |  |  |  | 10 | 16.2 | 5.4 |  | 7.63 | 11.59 | 0.86 | 678 | 175 |
|  |  |  |  | 15 | 21.0 | 5.3 |  | 7.61 | 11.47 | 0.85 | 680 | 175 |
|  |  | **Evita V600** | **CPAP** | 0 | 6.0 | 0.9 | 1.83 | 11.88 | 22.83 | 1.37 | 548 | 216 |
|  |  |  |  | 5 | 9.7 | 0.5 | 1.48 | 11.41 | 21.14 | 1.31 | 573 | 214 |
|  |  |  |  | 10 | 14.7 | 1.3 | 1.58 | 11.33 | 20.47 | 1.29 | 592 | 222 |
|  |  |  |  | 15 | 20.4 | 1.8 | 2.23 | 12.00 | 21.79 | 1.37 | 591 | 227 |
|  |  |  | **ATC** | 0 | 6.3 | 2.6 |  | 9.54 | 16.85 | 1.17 | 623 | 184 |
|  |  |  |  | 5 | 7.5 | 2.6 |  | 7.55 | 10.43 | 0.79 | 706 | 170 |
|  |  |  |  | 10 | 11.6 | 3.4 |  | 7.19 | 8.14 | 0.62 | 716 | 149 |
|  |  |  |  | 15 | 15.6 | 2.2 |  | 7.05 | 7.76 | 0.60 | 722 | 150 |
|  |  | **Elisa 800** | **CPAP** | 0 | 6.2 | 1.9 | 2.27 | 11.76 | 22.90 | 1.39 | 559 | 219 |
|  |  |  |  | 5 | 9.9 | -0.1 | 1.45 | 10.95 | 20.48 | 1.28 | 565 | 193 |
|  |  |  |  | 10 | 15.3 | 0.3 | 1.71 | 11.23 | 21.18 | 1.33 | 569 | 201 |
|  |  |  |  | 15 | 20.6 | 0.6 | 1.97 | 11.47 | 21.74 | 1.37 | 573 | 207 |
|  |  |  | **ATC** | 0 | 6.2 | 2.0 |  | 9.98 | 17.93 | 1.20 | 584 | 187 |
|  |  |  |  | 5 | 7.3 | 0.9 |  | 7.89 | 11.55 | 0.84 | 655 | 147 |
|  |  |  |  | 10 | 11.6 | 1.3 |  | 7.70 | 10.44 | 0.77 | 665 | 144 |
|  |  |  |  | 15 | 17.0 | 1.8 |  | 7.95 | 11.01 | 0.81 | 672 | 152 |
|  |  | **Hamilton C2** | **CPAP** | 0 | 6.9 | 0.5 | 4.37 | 13.89 | 28.47 | 1.51 | 488 | 237 |
|  |  |  |  | 5 | 10.3 | 0.4 | 3.26 | 12.98 | 25.42 | 1.39 | 513 | 234 |
|  |  |  |  | 10 | 15.3 | 0.5 | 3.21 | 12.95 | 25.23 | 1.40 | 519 | 233 |
|  |  |  |  | 15 | 20.3 | 0.5 | 3.19 | 12.95 | 25.23 | 1.41 | 523 | 233 |
|  |  |  | **ATC** | 0 | 7.2 | 0.9 |  | 11.81 | 23.13 | 1.37 | 533 | 214 |
|  |  |  |  | 5 | 8.6 | 0.4 |  | 9.19 | 15.45 | 1.02 | 604 | 187 |
|  |  |  |  | 10 | 12.6 | 0.8 |  | 8.89 | 13.94 | 0.94 | 619 | 184 |
|  |  |  |  | 15 | 17.6 | 0.3 |  | 8.95 | 13.83 | 0.94 | 623 | 180 |
|  |  | **Hamilton C6** | **CPAP** | 0 | 8.3 | 1.5 | 6.39 | 15.84 | 34.92 | 1.75 | 471 | 265 |
|  |  |  |  | 5 | 11.5 | 1.2 | 3.68 | 13.46 | 27.35 | 1.54 | 522 | 244 |
|  |  |  |  | 10 | 16.4 | 1.2 | 3.54 | 13.34 | 27.07 | 1.54 | 527 | 241 |
|  |  |  |  | 15 | 21.3 | 1.4 | 3.58 | 13.39 | 26.91 | 1.54 | 531 | 242 |
|  |  |  | **ATC** | 0 | 8.5 | 1.7 |  | 13.32 | 27.40 | 1.58 | 513 | 237 |
|  |  |  |  | 5 | 10.1 | 0.9 |  | 10.74 | 19.81 | 1.24 | 571 | 205 |
|  |  |  |  | 10 | 14.9 | 1.0 |  | 10.64 | 19.47 | 1.23 | 578 | 205 |
|  |  |  |  | 15 | 19.8 | 1.2 |  | 10.58 | 19.21 | 1.22 | 582 | 203 |
| **8** | **weak** | **TPiece** | **TPiece** | 0 | 0.0 |  |  | 4.47 | 4.80 | 0.15 | 324 | 188 |
|  |  | **Prototype** | **CPAP** | 0 | 0.0 | 0.2 | 0.43 | 4.25 | 4.60 | 0.15 | 342 | 194 |
|  |  |  |  | 5 | 5.0 | 0.2 | 0.45 | 4.25 | 4.43 | 0.15 | 344 | 192 |
|  |  |  |  | 10 | 10.0 | 0.2 | 0.45 | 4.27 | 4.55 | 0.15 | 346 | 194 |
|  |  |  | **ATC** | 0 | 0.0 | 0.1 |  | 1.19 | 1.05 | 0.05 | 457 | 157 |
|  |  |  |  | 5 | 5.0 | 0.1 |  | 1.23 | 0.99 | 0.05 | 456 | 157 |
|  |  |  |  | 10 | 10.0 | 0.1 |  | 1.26 | 1.06 | 0.05 | 455 | 156 |
|  |  | **EVITA4** | **CPAP** | 0 | 0.3 | 0.9 | 2.41 | 6.23 | 7.25 | 0.20 | 296 | 257 |
|  |  |  |  | 5 | 5.1 | 0.9 | 1.88 | 5.67 | 6.52 | 0.19 | 314 | 270 |
|  |  |  |  | 10 | 10.3 | 1.0 | 2.04 | 5.83 | 7.27 | 0.21 | 307 | 270 |
|  |  |  | **ATC** | 0 | 0.5 | 1.1 |  | 3.56 | 1.96 | 0.07 | 428 | 197 |
|  |  |  |  | 5 | 5.2 | 1.3 |  | 3.29 | 1.33 | 0.05 | 466 | 227 |
|  |  |  |  | 10 | 10.2 | 1.2 |  | 3.32 | 1.54 | 0.06 | 462 | 235 |
|  |  | **EVITAXL** | **CPAP** | 0 | 0.3 | 0.9 | 2.17 | 6.00 | 6.88 | 0.20 | 303 | 250 |
|  |  |  |  | 5 | 5.2 | 1.0 | 1.53 | 5.35 | 6.42 | 0.19 | 313 | 257 |
|  |  |  |  | 10 | 10.1 | 0.9 | 1.37 | 5.21 | 6.05 | 0.18 | 324 | 250 |
|  |  |  | **ATC** | 0 | 0.4 | 1.2 |  | 3.47 | 1.56 | 0.06 | 425 | 198 |
|  |  |  |  | 5 | 5.3 | 1.3 |  | 3.28 | 1.45 | 0.06 | 445 | 221 |
|  |  |  |  | 10 | 10.4 | 1.4 |  | 3.48 | 2.20 | 0.08 | 431 | 232 |
|  |  | **Evita V600** | **CPAP** | 0 | 0.4 | 0.7 | 1.74 | 5.40 | 7.02 | 0.21 | 310 | 252 |
|  |  |  |  | 5 | 4.9 | 0.6 | 1.50 | 4.97 | 5.71 | 0.18 | 335 | 251 |
|  |  |  |  | 10 | 9.8 | 0.5 | 1.55 | 4.89 | 5.67 | 0.18 | 341 | 257 |
|  |  |  | **ATC** | 0 | 0.4 | 0.6 |  | 4.76 | 5.67 | 0.18 | 341 | 241 |
|  |  |  |  | 5 | 4.9 | 0.8 |  | 3.78 | 4.13 | 0.15 | 386 | 257 |
|  |  |  |  | 10 | 9.8 | 0.9 |  | 3.75 | 3.89 | 0.15 | 399 | 245 |
|  |  | **Elisa 800** | **CPAP** | 0 | 0.4 | 0.9 | 1.97 | 5.57 | 6.51 | 0.19 | 313 | 222 |
|  |  |  |  | 5 | 5.0 | -0.1 | 0.70 | 4.50 | 5.27 | 0.17 | 333 | 211 |
|  |  |  |  | 10 | 9.9 | 0.1 | 0.94 | 4.65 | 5.16 | 0.17 | 339 | 212 |
|  |  |  | **ATC** | 0 | 0.4 | 1.0 |  | 4.90 | 4.92 | 0.16 | 346 | 204 |
|  |  |  |  | 5 | 4.8 | -0.1 |  | 3.46 | 3.64 | 0.14 | 384 | 194 |
|  |  |  |  | 10 | 9.8 | 0.2 |  | 3.68 | 3.70 | 0.14 | 390 | 203 |
|  |  | **Hamilton C2** | **CPAP** | 0 | 0.3 | 0.3 | 3.49 | 7.12 | 8.49 | 0.21 | 258 | 224 |
|  |  |  |  | 5 | 4.9 | 0.2 | 2.57 | 6.33 | 7.11 | 0.19 | 279 | 216 |
|  |  |  |  | 10 | 10.0 | 0.2 | 2.61 | 6.30 | 7.47 | 0.20 | 278 | 224 |
|  |  |  | **ATC** | 0 | 0.4 | 0.4 |  | 6.60 | 7.58 | 0.20 | 275 | 222 |
|  |  |  |  | 5 | 4.9 | 0.2 |  | 5.36 | 6.17 | 0.18 | 299 | 234 |
|  |  |  |  | 10 | 9.9 | 0.2 |  | 5.35 | 6.27 | 0.18 | 304 | 223 |
|  |  | **Hamilton C6** | **CPAP** | 0 | 0.7 | 1.0 | 5.57 | 8.93 | 12.68 | 0.27 | 234 | 323 |
|  |  |  |  | 5 | 5.0 | 0.8 | 2.94 | 6.57 | 7.79 | 0.21 | 292 | 277 |
|  |  |  |  | 10 | 10.1 | 0.9 | 2.96 | 6.58 | 7.90 | 0.22 | 293 | 276 |
|  |  |  | **ATC** | 0 | 0.7 | 1.1 |  | 7.99 | 11.21 | 0.26 | 258 | 316 |
|  |  |  |  | 5 | 5.0 | 0.8 |  | 5.85 | 6.86 | 0.20 | 318 | 275 |
|  |  |  |  | 10 | 10.1 | 0.9 |  | 5.92 | 7.09 | 0.21 | 316 | 275 |
| **8** | **moderate** | **TPiece** | **TPiece** | 0 | 0.0 |  |  | 5.81 | 5.45 | 0.23 | 414 | 137 |
|  |  | **Prototype** | **CPAP** | 0 | 0.0 | 0.4 | 0.49 | 5.52 | 5.16 | 0.23 | 428 | 147 |
|  |  |  |  | 5 | 5.0 | 0.4 | 0.52 | 5.54 | 5.19 | 0.23 | 434 | 144 |
|  |  |  |  | 10 | 10.0 | 0.3 | 0.53 | 5.54 | 5.23 | 0.23 | 430 | 147 |
|  |  |  |  | 15 | 15.0 | 0.3 | 0.52 | 5.52 | 5.17 | 0.23 | 433 | 142 |
|  |  |  | **ATC** | 0 | 0.0 | 0.1 |  | 1.42 | 0.94 | 0.05 | 504 | 102 |
|  |  |  |  | 5 | 5.0 | 0.1 |  | 1.46 | 0.94 | 0.05 | 503 | 102 |
|  |  |  |  | 10 | 9.9 | 0.2 |  | 1.46 | 0.92 | 0.05 | 500 | 106 |
|  |  |  |  | 15 | 15.0 | 0.1 |  | 1.42 | 0.94 | 0.05 | 506 | 103 |
|  |  | **EVITA4** | **CPAP** | 0 | 0.2 | 1.3 | 2.71 | 7.77 | 6.90 | 0.29 | 414 | 192 |
|  |  |  |  | 5 | 4.8 | 1.2 | 1.86 | 7.02 | 6.26 | 0.27 | 429 | 183 |
|  |  |  |  | 10 | 9.8 | 1.3 | 1.80 | 6.97 | 6.16 | 0.27 | 432 | 187 |
|  |  |  |  | 15 | 14.7 | 1.3 | 1.88 | 7.04 | 6.14 | 0.27 | 433 | 188 |
|  |  |  | **ATC** | 0 | 0.1 | 1.1 |  | 5.04 | 1.18 | 0.07 | 519 | 143 |
|  |  |  |  | 5 | 5.0 | 1.5 |  | 4.87 | 1.69 | 0.09 | 525 | 158 |
|  |  |  |  | 10 | 9.8 | 1.2 |  | 4.56 | 0.89 | 0.05 | 540 | 145 |
|  |  |  |  | 15 | 14.8 | 1.3 |  | 4.64 | 0.96 | 0.05 | 542 | 146 |
|  |  | **EVITAXL** | **CPAP** | 0 | 0.0 | 1.1 | 2.35 | 7.44 | 6.48 | 0.27 | 418 | 178 |
|  |  |  |  | 5 | 4.9 | 1.2 | 1.61 | 6.81 | 6.13 | 0.26 | 426 | 178 |
|  |  |  |  | 10 | 10.0 | 1.3 | 1.46 | 6.66 | 6.33 | 0.27 | 426 | 182 |
|  |  |  |  | 15 | 15.0 | 1.3 | 1.43 | 6.62 | 6.21 | 0.27 | 428 | 180 |
|  |  |  | **ATC** | 0 | 0.1 | 1.2 |  | 4.98 | 0.97 | 0.05 | 509 | 136 |
|  |  |  |  | 5 | 4.9 | 1.1 |  | 4.51 | 0.73 | 0.04 | 530 | 127 |
|  |  |  |  | 10 | 10.1 | 1.5 |  | 4.62 | 1.13 | 0.06 | 521 | 138 |
|  |  |  |  | 15 | 15.0 | 1.3 |  | 4.48 | 1.02 | 0.06 | 529 | 130 |
|  |  | **Evita V600** | **CPAP** | 0 | 0.3 | 0.9 | 1.64 | 6.67 | 6.49 | 0.28 | 423 | 181 |
|  |  |  |  | 5 | 4.9 | 0.9 | 1.44 | 6.26 | 6.09 | 0.27 | 425 | 179 |
|  |  |  |  | 10 | 9.6 | 0.9 | 1.69 | 6.24 | 5.69 | 0.25 | 436 | 171 |
|  |  |  |  | 15 | 14.7 | 1.0 | 1.92 | 6.23 | 6.01 | 0.27 | 430 | 178 |
|  |  |  | **ATC** | 0 | 0.3 | 0.9 |  | 5.98 | 4.85 | 0.23 | 459 | 165 |
|  |  |  |  | 5 | 4.7 | 1.1 |  | 4.97 | 3.70 | 0.19 | 490 | 152 |
|  |  |  |  | 10 | 9.8 | 1.2 |  | 5.00 | 4.22 | 0.21 | 485 | 165 |
|  |  |  |  | 15 | 15.1 | 1.3 |  | 5.00 | 4.79 | 0.23 | 481 | 180 |
|  |  | **Elisa 800** | **CPAP** | 0 | 0.3 | 1.3 | 2.12 | 6.87 | 6.42 | 0.28 | 431 | 179 |
|  |  |  |  | 5 | 5.0 | -0.1 | 1.11 | 5.99 | 5.96 | 0.26 | 419 | 151 |
|  |  |  |  | 10 | 9.8 | 0.2 | 1.44 | 6.23 | 5.66 | 0.25 | 437 | 152 |
|  |  |  |  | 15 | 14.7 | 0.4 | 1.70 | 6.39 | 5.56 | 0.25 | 445 | 157 |
|  |  |  | **ATC** | 0 | 0.2 | 1.3 |  | 6.19 | 4.78 | 0.23 | 451 | 165 |
|  |  |  |  | 5 | 4.9 | 0.0 |  | 4.56 | 4.06 | 0.20 | 467 | 129 |
|  |  |  |  | 10 | 9.8 | 0.3 |  | 4.80 | 3.96 | 0.20 | 465 | 135 |
|  |  |  |  | 15 | 14.7 | 0.6 |  | 5.04 | 3.85 | 0.19 | 479 | 139 |
|  |  | **Hamilton C2** | **CPAP** | 0 | 0.1 | 0.5 | 3.84 | 8.63 | 8.17 | 0.31 | 374 | 165 |
|  |  |  |  | 5 | 4.9 | 0.3 | 2.86 | 7.83 | 8.35 | 0.31 | 373 | 174 |
|  |  |  |  | 10 | 10.0 | 0.4 | 2.87 | 7.81 | 8.54 | 0.32 | 370 | 177 |
|  |  |  |  | 15 | 14.9 | 0.5 | 2.83 | 7.77 | 8.37 | 0.32 | 377 | 168 |
|  |  |  | **ATC** | 0 | 0.1 | 0.7 |  | 7.97 | 6.79 | 0.28 | 401 | 157 |
|  |  |  |  | 5 | 4.8 | 0.4 |  | 6.70 | 6.58 | 0.27 | 405 | 161 |
|  |  |  |  | 10 | 9.8 | 0.4 |  | 6.62 | 6.66 | 0.27 | 404 | 161 |
|  |  |  |  | 15 | 14.7 | 0.4 |  | 6.61 | 6.57 | 0.27 | 408 | 160 |
|  |  | **Hamilton C6** | **CPAP** | 0 | 0.4 | 1.6 | 5.95 | 10.45 | 10.90 | 0.39 | 362 | 232 |
|  |  |  |  | 5 | 4.7 | 1.2 | 3.24 | 8.11 | 7.40 | 0.30 | 411 | 197 |
|  |  |  |  | 10 | 9.7 | 1.2 | 3.06 | 7.98 | 7.28 | 0.30 | 413 | 198 |
|  |  |  |  | 15 | 14.8 | 1.4 | 3.21 | 8.10 | 7.67 | 0.31 | 408 | 205 |
|  |  |  | **ATC** | 0 | 0.4 | 1.5 |  | 9.08 | 8.33 | 0.34 | 405 | 222 |
|  |  |  |  | 5 | 4.8 | 1.2 |  | 7.07 | 5.87 | 0.26 | 436 | 184 |
|  |  |  |  | 10 | 9.8 | 1.1 |  | 6.98 | 5.92 | 0.26 | 439 | 188 |
|  |  |  |  | 15 | 14.9 | 1.3 |  | 7.06 | 6.42 | 0.28 | 422 | 197 |
| **8** | **strong** | **TPiece** | **TPiece** | 0 | 2.6 |  |  | 7.73 | 12.37 | 0.80 | 577 | 151 |
|  |  | **Prototype** | **CPAP** | 0 | 2.4 | 0.4 | 0.43 | 7.28 | 11.21 | 0.78 | 627 | 154 |
|  |  |  |  | 5 | 7.3 | 0.5 | 0.45 | 7.25 | 10.90 | 0.77 | 641 | 150 |
|  |  |  |  | 10 | 12.2 | 0.5 | 0.47 | 7.29 | 10.76 | 0.78 | 653 | 149 |
|  |  |  |  | 15 | 17.2 | 0.4 | 0.46 | 7.30 | 10.72 | 0.78 | 655 | 147 |
|  |  |  | **ATC** | 0 | -0.8 | 0.1 |  | 1.36 | 0.32 | 0.03 | 691 | 25 |
|  |  |  |  | 5 | 4.1 | 0.1 |  | 1.37 | 0.33 | 0.03 | 694 | 21 |
|  |  |  |  | 10 | 9.1 | 0.1 |  | 1.42 | 0.34 | 0.03 | 699 | 18 |
|  |  |  |  | 15 | 13.9 | 0.1 |  | 1.53 | 0.39 | 0.04 | 704 | 14 |
|  |  | **EVITA4** | **CPAP** | 0 | 5.1 | 1.4 | 2.89 | 10.03 | 18.10 | 1.14 | 566 | 194 |
|  |  |  |  | 5 | 8.8 | 1.6 | 2.19 | 9.37 | 15.80 | 1.03 | 594 | 188 |
|  |  |  |  | 10 | 13.6 | 1.5 | 2.01 | 9.21 | 15.50 | 1.02 | 599 | 185 |
|  |  |  |  | 15 | 18.5 | 1.5 | 1.93 | 9.13 | 15.25 | 1.01 | 604 | 184 |
|  |  |  | **ATC** | 0 | 5.1 | 3.9 |  | 6.60 | 9.46 | 0.69 | 649 | 146 |
|  |  |  |  | 5 | 9.0 | 3.9 |  | 6.15 | 8.02 | 0.60 | 673 | 142 |
|  |  |  |  | 10 | 13.6 | 3.4 |  | 5.97 | 7.55 | 0.56 | 671 | 142 |
|  |  |  |  | 15 | 18.5 | 3.6 |  | 5.75 | 7.05 | 0.54 | 686 | 135 |
|  |  | **EVITAXL** | **CPAP** | 0 | 4.7 | 1.3 | 2.63 | 9.75 | 17.33 | 1.10 | 571 | 190 |
|  |  |  |  | 5 | 8.5 | 1.6 | 2.08 | 9.22 | 15.05 | 0.99 | 598 | 181 |
|  |  |  |  | 10 | 13.3 | 1.6 | 1.89 | 9.04 | 14.66 | 0.97 | 602 | 177 |
|  |  |  |  | 15 | 18.3 | 1.6 | 1.83 | 8.95 | 14.49 | 0.97 | 607 | 176 |
|  |  |  | **ATC** | 0 | 4.9 | 3.3 |  | 6.36 | 9.33 | 0.68 | 641 | 145 |
|  |  |  |  | 5 | 8.8 | 3.6 |  | 5.83 | 7.28 | 0.54 | 672 | 135 |
|  |  |  |  | 10 | 13.8 | 3.6 |  | 5.62 | 7.14 | 0.53 | 674 | 133 |
|  |  |  |  | 15 | 18.7 | 3.6 |  | 5.59 | 7.05 | 0.53 | 680 | 131 |
|  |  | **Evita V600** | **CPAP** | 0 | 4.1 | 0.8 | 1.91 | 9.02 | 15.44 | 1.02 | 592 | 174 |
|  |  |  |  | 5 | 7.4 | 0.7 | 1.53 | 8.56 | 13.30 | 0.91 | 622 | 172 |
|  |  |  |  | 10 | 13.1 | 1.8 | 1.97 | 8.91 | 13.80 | 0.95 | 635 | 182 |
|  |  |  |  | 15 | 17.5 | 1.5 | 1.74 | 8.51 | 12.77 | 0.90 | 649 | 181 |
|  |  |  | **ATC** | 0 | 4.2 | 1.4 |  | 7.24 | 11.35 | 0.83 | 639 | 144 |
|  |  |  |  | 5 | 5.4 | 1.7 |  | 5.86 | 6.06 | 0.47 | 698 | 119 |
|  |  |  |  | 10 | 10.0 | 1.3 |  | 5.66 | 5.54 | 0.43 | 702 | 124 |
|  |  |  |  | 15 | 14.6 | 1.6 |  | 5.59 | 4.88 | 0.38 | 719 | 122 |
|  |  | **Elisa 800** | **CPAP** | 0 | 4.1 | 2.0 | 2.30 | 8.95 | 15.14 | 1.02 | 605 | 178 |
|  |  |  |  | 5 | 7.9 | 0.1 | 1.59 | 8.24 | 13.36 | 0.92 | 607 | 153 |
|  |  |  |  | 10 | 13.3 | 0.4 | 1.86 | 8.52 | 14.01 | 0.96 | 612 | 162 |
|  |  |  |  | 15 | 18.6 | 0.8 | 2.11 | 8.76 | 14.50 | 1.00 | 616 | 167 |
|  |  |  | **ATC** | 0 | 4.0 | 2.1 |  | 7.78 | 12.11 | 0.86 | 619 | 155 |
|  |  |  |  | 5 | 5.3 | 0.7 |  | 5.84 | 6.34 | 0.48 | 672 | 105 |
|  |  |  |  | 10 | 10.6 | 1.1 |  | 6.07 | 6.88 | 0.52 | 675 | 112 |
|  |  |  |  | 15 | 15.9 | 1.5 |  | 6.32 | 7.34 | 0.56 | 682 | 121 |
|  |  | **Hamilton C2** | **CPAP** | 0 | 5.4 | 0.6 | 4.43 | 11.25 | 21.18 | 1.24 | 532 | 206 |
|  |  |  |  | 5 | 8.6 | 0.5 | 3.21 | 10.19 | 17.86 | 1.09 | 561 | 196 |
|  |  |  |  | 10 | 13.5 | 0.7 | 3.17 | 10.18 | 17.67 | 1.09 | 567 | 197 |
|  |  |  |  | 15 | 18.5 | 0.7 | 3.20 | 10.19 | 17.59 | 1.09 | 572 | 197 |
|  |  |  | **ATC** | 0 | 5.6 | 0.8 |  | 9.94 | 18.05 | 1.14 | 560 | 190 |
|  |  |  |  | 5 | 7.7 | 0.4 |  | 8.35 | 13.37 | 0.89 | 601 | 173 |
|  |  |  |  | 10 | 12.4 | 0.4 |  | 8.30 | 13.05 | 0.87 | 611 | 171 |
|  |  |  |  | 15 | 17.3 | 0.1 |  | 8.25 | 12.65 | 0.86 | 619 | 168 |
|  |  | **Hamilton C6** | **CPAP** | 0 | 6.8 | 1.6 | 6.38 | 13.11 | 26.94 | 1.51 | 515 | 234 |
|  |  |  |  | 5 | 9.7 | 1.1 | 3.73 | 10.69 | 19.71 | 1.24 | 567 | 205 |
|  |  |  |  | 10 | 14.6 | 1.3 | 3.66 | 10.65 | 19.45 | 1.23 | 573 | 203 |
|  |  |  |  | 15 | 19.5 | 1.4 | 3.65 | 10.65 | 19.32 | 1.23 | 577 | 203 |
|  |  |  | **ATC** | 0 | 6.9 | 1.5 |  | 11.43 | 22.12 | 1.37 | 543 | 206 |
|  |  |  |  | 5 | 9.1 | 0.9 |  | 9.32 | 15.64 | 1.05 | 597 | 182 |
|  |  |  |  | 10 | 13.6 | 0.9 |  | 9.10 | 15.23 | 1.03 | 603 | 177 |
|  |  |  |  | 15 | 18.5 | 1.0 |  | 9.06 | 15.14 | 1.03 | 604 | 176 |

**Table S2:** Effectiveness of tube compensation. Percentage of tube compensation based on the percent reduction of R_eff,trach_, inspiratory PTP or inspiratory WOB_add_ under ATC relative to the corresponding values measured during a T-piece trial (from Table S1)

| **ETT** | **Ventilator** | **PEEP** | **Effort** |  |  |  |  |  |  |  |  |
|---|---|---|---|---|---|---|---|---|---|---|---|
|  |  |  | **weak** |  |  | **moderate** |  |  | **strong** |  |  |
|  |  |  | **R<sub>eff</sub>** | **PTP** | **WOB** | **R<sub>eff</sub>** | **PTP** | **WOB** | **R<sub>eff</sub>** | **PTP** | **WOB** |
| *mm* |  | *mbar* | *%* | *%* | *%* | *%* | *%* | *%* | *%* | *%* | *%* |
| **7** | **Prototype** | 0 | 78 | 89 | 81 | 78 | 88 | 82 | 88 | 100 | 100 |
|  |  | 5 | 78 | 88 | 80 | 78 | 88 | 82 | 88 | 100 | 100 |
|  |  | 10 | 78 | 91 | 85 | 78 | 89 | 83 | 86 | 99 | 98 |
|  |  | 15 |  |  |  | 78 | 90 | 84 | 86 | 99 | 98 |
|  | **EVITA4** | 0 | 27 | 54 | 38 | 20 | 74 | 63 | 18 | 23 | 8 |
|  |  | 5 | 26 | 58 | 47 | 25 | 79 | 70 | 17 | 25 | 14 |
|  |  | 10 | 28 | 52 | 38 | 25 | 74 | 63 | 21 | 33 | 17 |
|  |  | 15 |  |  |  | 19 | 62 | 52 | 23 | 31 | 15 |
|  | **EVITAXL** | 0 | 24 | 52 | 37 | 22 | 76 | 66 | 22 | 29 | 14 |
|  |  | 5 | 34 | 70 | 57 | 25 | 77 | 68 | 26 | 37 | 22 |
|  |  | 10 | 36 | 71 | 58 | 27 | 78 | 68 | 28 | 40 | 25 |
|  |  | 15 |  |  |  | 28 | 80 | 71 | 29 | 41 | 26 |
|  | **EvitaV600** | 0 | -1 | -15 | -20 | 5 | 21 | 6 | 11 | 13 | -3 |
|  |  | 5 | 21 | 21 | 4 | 23 | 39 | 22 | 29 | 46 | 31 |
|  |  | 10 | 23 | 25 | 6 | 24 | 41 | 24 | 33 | 58 | 45 |
|  |  | 15 |  |  |  | 25 | 28 | 10 | 34 | 60 | 48 |
|  | **Elisa 800** | 0 | 0 | 2 | -7 | 2 | 19 | 6 | 6 | 8 | -5 |
|  |  | 5 | 27 | 27 | 10 | 27 | 28 | 13 | 26 | 41 | 27 |
|  |  | 10 | 23 | 27 | 10 | 25 | 31 | 16 | 28 | 46 | 33 |
|  |  | 15 |  |  |  | 22 | 33 | 18 | 26 | 43 | 29 |
|  | **HamiltonC2** | 0 | -27 | -37 | -21 | -17 | -6 | -7 | -11 | -19 | -20 |
|  |  | 5 | 0 | -6 | -6 | 6 | 1 | -3 | 14 | 21 | 11 |
|  |  | 10 | 1 | -6 | -6 | 7 | 0 | -4 | 17 | 28 | 18 |
|  |  | 15 |  |  |  | 5 | 3 | -3 | 16 | 29 | 18 |
|  | **HamiltonC6** | 0 | -55 | -98 | -49 | -35 | -33 | -30 | -25 | -41 | -38 |
|  |  | 5 | -16 | -27 | -22 | -9 | -3 | -8 | -1 | -2 | -9 |
|  |  | 10 | -17 | -32 | -24 | -5 | -2 | -8 | 0 | 0 | -8 |
|  |  | 15 |  |  |  | -6 | -8 | -13 | 1 | 1 | -7 |
| **8** | **Prototype** | 0 | 73 | 78 | 68 | 76 | 83 | 77 | 82 | 97 | 96 |
|  |  | 5 | 72 | 79 | 70 | 75 | 83 | 77 | 82 | 97 | 96 |
|  |  | 10 | 72 | 78 | 68 | 75 | 83 | 77 | 82 | 97 | 96 |
|  |  | 15 |  |  |  | 76 | 83 | 76 | 80 | 97 | 95 |
|  | **EVITA4** | 0 | 20 | 59 | 51 | 13 | 78 | 72 | 15 | 24 | 14 |
|  |  | 5 | 27 | 72 | 66 | 16 | 69 | 63 | 20 | 35 | 25 |
|  |  | 10 | 26 | 68 | 60 | 22 | 84 | 79 | 23 | 39 | 30 |
|  |  | 15 |  |  |  | 20 | 82 | 77 | 26 | 43 | 33 |
|  | **EVITAXL** | 0 | 22 | 68 | 60 | 14 | 82 | 77 | 18 | 25 | 15 |
|  |  | 5 | 27 | 70 | 62 | 22 | 87 | 82 | 25 | 41 | 32 |
|  |  | 10 | 22 | 54 | 45 | 21 | 79 | 73 | 27 | 42 | 33 |
|  |  | 15 |  |  |  | 23 | 81 | 75 | 28 | 43 | 33 |
|  | **EvitaV600** | 0 | -6 | -18 | -20 | -3 | 11 | 1 | 6 | 8 | -4 |
|  |  | 5 | 16 | 14 | 2 | 14 | 32 | 19 | 24 | 51 | 41 |
|  |  | 10 | 16 | 19 | 5 | 14 | 22 | 9 | 27 | 55 | 46 |
|  |  | 15 |  |  |  | 14 | 12 | -1 | 28 | 61 | 52 |
|  | **Elisa 800** | 0 | -10 | -3 | -8 | -7 | 12 | 2 | -1 | 2 | -7 |
|  |  | 5 | 23 | 24 | 11 | 22 | 25 | 14 | 24 | 49 | 40 |
|  |  | 10 | 18 | 23 | 10 | 17 | 27 | 15 | 21 | 44 | 34 |
|  |  | 15 |  |  |  | 13 | 29 | 17 | 18 | 41 | 30 |
|  | **HamiltonC2** | 0 | -47 | -58 | -33 | -37 | -25 | -19 | -29 | -46 | -43 |
|  |  | 5 | -20 | -29 | -18 | -15 | -21 | -17 | -8 | -8 | -11 |
|  |  | 10 | -20 | -31 | -19 | -14 | -22 | -18 | -7 | -5 | -10 |
|  |  | 15 |  |  |  | -14 | -21 | -18 | -7 | -2 | -8 |
|  | **HamiltonC6** | 0 | -79 | -134 | -70 | -56 | -53 | -46 | -48 | -79 | -72 |
|  |  | 5 | -31 | -43 | -32 | -22 | -8 | -11 | -21 | -26 | -31 |
|  |  | 10 | -32 | -48 | -35 | -20 | -9 | -13 | -18 | -23 | -29 |
|  |  | 15 |  |  |  | -21 | -18 | -20 | -17 | -22 | -29 |

**Table S3:** Effectiveness of tube compensation. Percentage of tube compensation under ATC relative to the corresponding values measured under CPAP for the same ventilator (from Table S1). Similar values between ventilators do not indicate a similar reduction in total load or work.

| **ETT** | **Ventilator** | **PEEP** | **Effort** |  |  |  |  |  |  |  |  |
|---|---|---|---|---|---|---|---|---|---|---|---|
|  |  |  | **weak** |  |  | **moderate** |  |  | **strong** |  |  |
|  |  |  | **R<sub>eff</sub>** | **PTP** | **WOB** | **R<sub>eff</sub>** | **PTP** | **WOB** | **R<sub>eff</sub>** | **PTP** | **WOB** |
| *mm* |  | *mbar* | *%* | *%* | *%* | *%* | *%* | *%* | *%* | *%* | *%* |
| **7** | **Prototype** | 0 | 77 | 88 | 80 | 77 | 88 | 82 | 88 | 100 | 100 |
|  |  | 5 | 77 | 88 | 80 | 77 | 88 | 82 | 87 | 100 | 100 |
|  |  | 10 | 77 | 91 | 85 | 77 | 89 | 83 | 86 | 99 | 99 |
|  |  | 15 |  |  |  | 77 | 90 | 84 | 85 | 99 | 98 |
|  | **EVITA4** | 0 | 43 | 67 | 50 | 35 | 78 | 68 | 32 | 42 | 27 |
|  |  | 5 | 38 | 69 | 56 | 35 | 81 | 73 | 27 | 37 | 27 |
|  |  | 10 | 41 | 66 | 51 | 34 | 77 | 68 | 30 | 45 | 31 |
|  |  | 15 |  |  |  | 30 | 67 | 59 | 31 | 41 | 28 |
|  | **EVITAXL** | 0 | 39 | 65 | 49 | 35 | 80 | 71 | 33 | 43 | 30 |
|  |  | 5 | 42 | 78 | 65 | 33 | 80 | 71 | 34 | 45 | 32 |
|  |  | 10 | 43 | 75 | 63 | 33 | 80 | 72 | 36 | 47 | 34 |
|  |  | 15 |  |  |  | 34 | 83 | 75 | 35 | 48 | 36 |
|  | **EvitaV600** | 0 | 13 | 18 | 7 | 14 | 31 | 19 | 20 | 26 | 14 |
|  |  | 5 | 30 | 36 | 19 | 27 | 44 | 31 | 34 | 51 | 40 |
|  |  | 10 | 30 | 39 | 22 | 27 | 44 | 31 | 37 | 60 | 52 |
|  |  | 15 |  |  |  | 28 | 32 | 19 | 41 | 64 | 56 |
|  | **Elisa 800** | 0 | 16 | 26 | 13 | 13 | 30 | 20 | 15 | 22 | 14 |
|  |  | 5 | 27 | 33 | 18 | 28 | 34 | 23 | 28 | 44 | 35 |
|  |  | 10 | 26 | 31 | 17 | 27 | 34 | 23 | 31 | 51 | 42 |
|  |  | 15 |  |  |  | 25 | 34 | 24 | 31 | 49 | 41 |
|  | **HamiltonC2** | 0 | 11 | 12 | 3 | 12 | 23 | 12 | 15 | 19 | 9 |
|  |  | 5 | 23 | 25 | 11 | 26 | 28 | 15 | 29 | 39 | 27 |
|  |  | 10 | 24 | 26 | 12 | 26 | 29 | 16 | 31 | 45 | 33 |
|  |  | 15 |  |  |  | 24 | 31 | 18 | 31 | 45 | 34 |
|  | **HamiltonC6** | 0 | 17 | 22 | 9 | 17 | 30 | 16 | 16 | 22 | 10 |
|  |  | 5 | 19 | 23 | 11 | 16 | 25 | 17 | 20 | 28 | 19 |
|  |  | 10 | 18 | 22 | 10 | 18 | 24 | 15 | 20 | 28 | 20 |
|  |  | 15 |  |  |  | 18 | 21 | 13 | 21 | 29 | 21 |
| **8** | **Prototype** | 0 | 72 | 77 | 68 | 74 | 82 | 77 | 81 | 97 | 96 |
|  |  | 5 | 71 | 78 | 69 | 74 | 82 | 77 | 81 | 97 | 96 |
|  |  | 10 | 71 | 77 | 68 | 74 | 83 | 77 | 81 | 97 | 96 |
|  |  | 15 |  |  |  | 74 | 82 | 77 | 79 | 96 | 95 |
|  | **EVITA4** | 0 | 43 | 73 | 63 | 35 | 83 | 77 | 34 | 48 | 40 |
|  |  | 5 | 42 | 80 | 73 | 31 | 73 | 69 | 34 | 49 | 42 |
|  |  | 10 | 43 | 79 | 71 | 35 | 85 | 82 | 35 | 51 | 45 |
|  |  | 15 |  |  |  | 34 | 84 | 80 | 37 | 54 | 47 |
|  | **EVITAXL** | 0 | 42 | 77 | 70 | 33 | 85 | 80 | 35 | 46 | 38 |
|  |  | 5 | 39 | 77 | 69 | 34 | 88 | 84 | 37 | 52 | 45 |
|  |  | 10 | 33 | 64 | 55 | 31 | 82 | 77 | 38 | 51 | 45 |
|  |  | 15 |  |  |  | 32 | 84 | 79 | 38 | 51 | 45 |
|  | **EvitaV600** | 0 | 12 | 19 | 11 | 10 | 25 | 17 | 20 | 26 | 19 |
|  |  | 5 | 24 | 28 | 17 | 21 | 39 | 30 | 32 | 54 | 48 |
|  |  | 10 | 23 | 31 | 20 | 20 | 26 | 17 | 36 | 60 | 55 |
|  |  | 15 |  |  |  | 20 | 20 | 12 | 34 | 62 | 57 |
|  | **Elisa 800** | 0 | 12 | 24 | 15 | 10 | 25 | 19 | 13 | 20 | 16 |
|  |  | 5 | 23 | 31 | 21 | 24 | 32 | 24 | 29 | 53 | 47 |
|  |  | 10 | 21 | 28 | 19 | 23 | 30 | 23 | 29 | 51 | 45 |
|  |  | 15 |  |  |  | 21 | 31 | 23 | 28 | 49 | 44 |
|  | **HamiltonC2** | 0 | 7 | 11 | 4 | 8 | 17 | 10 | 12 | 15 | 8 |
|  |  | 5 | 15 | 13 | 5 | 14 | 21 | 13 | 18 | 25 | 19 |
|  |  | 10 | 15 | 16 | 8 | 15 | 22 | 14 | 18 | 26 | 20 |
|  |  | 15 |  |  |  | 15 | 22 | 14 | 19 | 28 | 21 |
|  | **HamiltonC6** | 0 | 11 | 12 | 4 | 13 | 24 | 13 | 13 | 18 | 9 |
|  |  | 5 | 11 | 12 | 5 | 13 | 21 | 15 | 13 | 21 | 15 |
|  |  | 10 | 10 | 10 | 4 | 13 | 19 | 13 | 15 | 22 | 17 |
|  |  | 15 |  |  |  | 13 | 16 | 11 | 15 | 22 | 17 |

